# Modeling the transmission of the SARS-CoV-2 delta variant in a partially vaccinated population

**DOI:** 10.1101/2021.09.23.21264032

**Authors:** Ugo Avila-Ponce de León, Eric Avila-Vales, Kuan-lin Huang

## Abstract

In a population with ongoing vaccination, the trajectory of a pandemic is determined by how the virus spreads in unvaccinated and vaccinated individuals that exhibit distinct transmission dynamics based on different levels of natural and vaccine-induced immunity. We developed a mathematical model that considers both subpopulations and immunity parameters including vaccination rates, vaccine effectiveness, and a gradual loss of protection. The model forecasted the spread of the SARS-CoV-2 delta variant in the US under varied transmission and vaccination rates. We further obtained the control reproduction number and conducted sensitivity analyses to determine how each parameter may affect virus transmission. Our results show that a combination of strengthening vaccine-induced immunity and preventative behavioral measures will likely be required to deaccelerate the rise of infectious SARS-CoV-2 variants.

**One-Sentence Summary:** Mathematical models considering vaccinated and unvaccinated individuals help forecast and manage the spread of new SARS-CoV-2 variants.

## Main Text

Severe acute respiratory syndrome coronavirus 2 (SARS-CoV-2) was first reported in late 2019 and has since amounted to over 200 million confirmed cases of COVID-19 around the globe (*1*). SARS-CoV-2 is an RNA virus that evolves through mutagenesis, and multiple new variants have emerged throughout the ongoing COVID-19 pandemic. Based on their varied levels of infectiousness, lethality, and response to the vaccine, the most threatening SARS-CoV-2 variants are identified as variants of concern (VOCs) (*2*). Succeeding the alpha variant (B.1.1.7 under the Pango lineage) that gained prevalence in the first half of 2021, the delta variant (B.1.617.2, AY.1, AY.2, AY.3) has become the most prominent strain among the sequenced variants in many countries by September 2021 *(3,4)*. While most of the approved vaccines still demonstrate effectiveness against SARS-CoV-2, the delta variant’s enhanced transmission rate and resistance to vaccines (*5*) imply that the future trajectory of the COVID-19 pandemic will depend on several parameters associated with vaccine-induced immunity. For example, a vaccine is considered leaky if it reduces but does not eliminate the possibility of infection in vaccinated individuals (<100% of vaccine effectiveness). As time passes, a higher fraction of the population may become vaccinated, but vaccine-induced immunity also wanes at a given rate. These parameters directly influence disease transmission in a population with an ongoing vaccination program, which describes the state of most countries in the current COVID-19 pandemic.

Multiple mathematical models have been developed to project the dynamics of infectious diseases, many of which extend upon the Susceptible-Infected-Recovered (SIR) compartmental model. Recent model innovations, such as accounting for secondary infections (*6*) or population sub-structures (*7*) have improved disease projections and prioritization of vaccine regimes. However, most models do not consider the distinct spread dynamics in the unvaccinated and vaccinated subpopulations, especially considering characteristics of the delta variant. A model for the partially-vaccinated population is required to understand the ongoing SARS-CoV-2 pandemic and will likely apply to the development of vaccination programs for future infectious viruses.

### Modelling the SARS-CoV-2 delta variant spread in a partially vaccinated population

We developed a compartmental differential equation to understand the dynamics of the spread of the SARS-CoV-2 delta variant under one vaccination program of a two-dose scheme (*8*), i.e., the BNT162b2 and ChAdOx1nCoV-19 vaccines. The mathematical model contains two separate sets of equations for the variant that affects both unvaccinated and vaccinated individuals. Fig 1A shows the flow between the compartmentalized subpopulations, Table S1 describes the parameters used in our mathematical model, and the Supplementary Texts provide an in-depth derivation of the mathematical. The SPFEIARD model evaluates the dynamics of eleven subpopulations at any given time t. *S*(*t*) denotes the susceptible population, *P*(*t*) denotes individuals with partial immunity provided by one dose of the vaccine, *F*(*t*) represents individuals with full immunity acquired by two doses of the vaccine. *E*(*t*) indicates exposed individuals that are unvaccinated. *I*(*t*) unvaccinated, infected individuals showing COVID-19 symptoms, *A*(*t*) unvaccinated, infected individuals that are asymptomatic. *E*_*B*_(*t*) denotes vaccinated (≥ 1) dose, exposed individuals. *I*_*B*_(*t*) denotes vaccinated (≥ 1) yet infected individuals, also considered as “breakthrough cases”. *A*_*B*_(*t*) represents vaccinated (≥ 1), infected individuals that are asymptomatic. *R*(*t*) denotes the population recovered from the disease regardless of their vaccination status. Finally, *D*(*t*) represents the deceased COVID-19 patients, regardless of their vaccination status.

**Figure 1.**
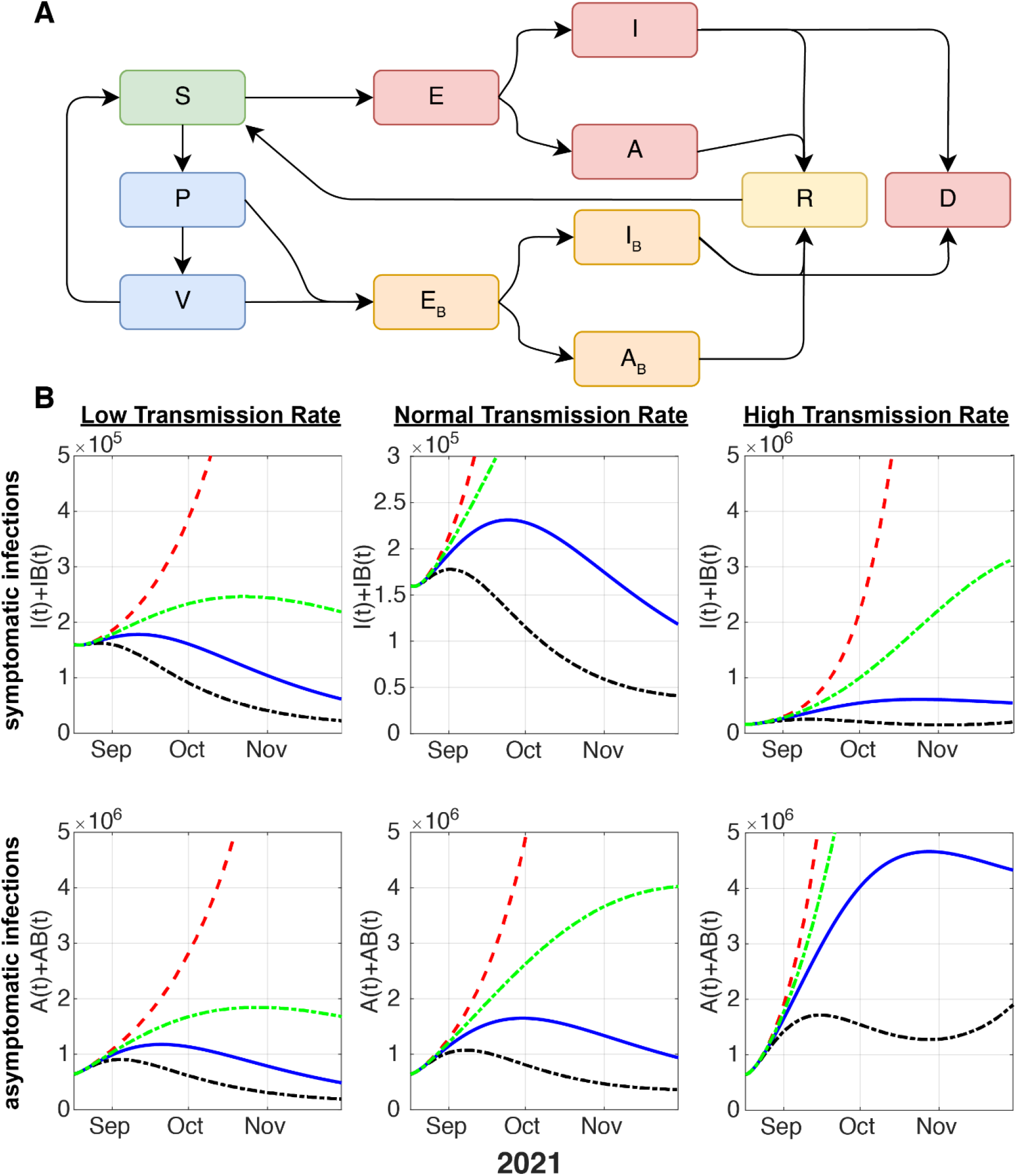
Schematic of the model considering vaccinated vs. unvaccinated individuals and projection of newly infected symptomatic and asymptomatic cases. **(A)** Flow dynamics of the mathematical model. S: susceptible, P: partial immunity (one vaccine dose), V: full immunity (two vaccine doses). E: unvaccinated, exposed. I: unvaccinated, infected and symptomatic, A: unvaccinated, infected, asymptomatic. E_B_: vaccinated (≥ 1 dose), exposed. I_B_: vaccinated (≥ 1 dose), infected, and symptomatic (“breakthrough cases”). A_B_: vaccinated (≥ 1 dose), infected and asymptomatic. R: recovered. D: deceased. **(B)** Modelled dynamics of the new infections that are symptomatic (top row) and asymptomatic (bottom row) from the SARS-CoV-2 delta variant considering different vaccination rates. The red line presents zero vaccination; the green line represents a 50% decrease in VR; the blue dotted line means baseline VR and the black dotted line denotes 200% VR.

We simulated immunity acquired by one dose or two doses of the BNT162b2 vaccine (the first US FDA-approved COVID-19 vaccine) against the delta variant, where the parameters were obtained from real-world data *(9)*. Vaccine effectiveness for the delta variant for only one dose for symptomatic COVID-19 is 0.35 (95% confidence interval (CI), 0.22-0.4). For two doses, the effectiveness is 0.59 (CI, 0.5-0.78). For asymptomatic infections, the effectiveness for one dose is 0.3 (CI, 0.22-0.4) and for two-dose is 0.49 (0.45-0.64). The all-or-nothing protection, which means people who received the vaccine but still developed COVID-19, is 0.0862 (CI, 0.0689,0.10344). The waning immunity parameter *α* was assumed herein by vaccine-induced immunity being worn off after six months and represented a conservative estimate that may apply for a variant with high resistance to vaccine-induced immunity. We also obtained the transmission rates for symptomatic and asymptomatic infections by fitting the parameters using data of daily infections and deaths provided by the repository developed by the Johns Hopkins University (1).

Given that in real-world situations, the vaccination rate is rarely a linear function defined by a constant daily dose, our model incorporates a piece-wise function of vaccination rate *ρ*(*t*) based on the applied daily doses from December 20th, 2020 to August 17th, 2021 (8). The projection of this baseline vaccination rate (VR) estimates that the US, by August 2021, has ∼45% of the population with one dose (Fig S1A), and ∼40% individuals with two doses of the vaccine (Fig S1B), and by October ∼60% with one dose and ∼50% with two doses (Fig S1C and D). The real-world VR may vary due to a wide range of factors, and thus we simulated different VRs from mid-August of 2021 based on this VR function in subsequent models.

### Spread of the delta variant under different transmission and vaccination rates

Using this model, we evaluate how different vaccine rates might affect the spread of the SARS-CoV-2 delta variant. We further considered low, normal, and high transmission rates, which could reflect different implementation levels of non-pharmaceutical strategies (NPI). Fig 1B shows the projections of new SARS-CoV-2 symptomatic and asymptomatic infections, which included the modelled solutions assuming no vaccination (red dotted line), 50% decrease of VR (green dotted line), baseline VR (blue solid line) and 200% VR (black dotted line). Regardless of the transmission rate, case counts would rise exponentially in a hypothetical US population with zero vaccination. Given a low transmission rate, new infections would increase to a peak of 2.45 * 10e5 symptomatic cases per day given 50% VR, 1.85 *10e5 given baseline VR, and 1.62 *10e5 given 200% VR, before tapering down in October or November. Asymptomatic individuals behave in the same manner as symptomatic (Fig 1B). Under a normal transmission rate, a US population with 50% VR would rise to a peak of 5.0 * 10e5 symptomatic cases and 4.00 * 10e6 asymptomatic cases per day. In comparison, the baseline and 200% VR population would have a more moderate pandemic, peaking at 2.35 * 10e5 and 1.86 * 10e6 symptomatic cases per day, respectively (Fig 1B). Under a high transmission rate, new infection counts would be significantly higher even under a baseline VR, reaching a peak of up to 0.89* 10e5 symptomatic cases and 4.76 * 10e6 asymptomatic cases per day. A 200% VR would be required to control the pandemic in this high transmission rate population, where the peak of new daily infections would still rise to 0.25* 10e5 symptomatic cases and 1.85 * 10e6 asymptomatic cases (Fig 1B).

### Vaccine effectiveness and new infections in vaccinated vs. unvaccinated individuals

Vaccinated individuals have consistently shown a significantly lower rate of contracting COVID-19. However, since the emergence of SARS-CoV-2 variants capable of immune evasion, an elevated number of breakthrough cases have been reported. We dissected the modelled results to determine new infections that would arise from vaccinated vs. unvaccinated subpopulations. In addition to using the estimate of vaccine effectiveness, we also constructed models of low and high vaccine effectiveness based on the upper and lower bound of the 95% confidence intervals *(9)*. Low vaccine effectiveness is modelled as 0.22 for one-dose vaccinated individuals for either symptomatic and asymptomatic infections, and for fully vaccinated individuals, 0.5 for asymptomatic and 0.45 for asymptomatic infections. High vaccine effectiveness is modelled as 0.4 for one-dose vaccinated individuals for either symptomatic and asymptomatic infections, and for fully vaccinated individuals, 0.78 for symptomatic and 0.64 for asymptomatic infections.

The models showed that, even considering the reduced vaccine effectiveness against the delta variant, the number of symptomatic infections contributed by the unvaccinated individuals is generally an order of magnitude (∼ 10x) higher than that from the vaccinated individuals under all scenarios (Fig 2). Given 50% VR, new symptomatic infections will continue to rise in unvaccinated individuals and surpass 5*10e5 cases by December 2021 regardless of vaccine effectiveness. In the case of baseline or 200% VR, symptomatic infections of unvaccinated individuals will peak at the end or beginning of September 2021, respectively, and continue to decrease. Notably, due to the higher number of individuals becoming vaccinated under baseline or 200% VR scenarios, new, symptomatic infections from vaccinated individuals (“breakthrough cases”) were projected to rise in October 2021 if the vaccine effectiveness is at low or baseline levels. If the vaccine effectiveness remains at a high level, the number of breakthrough cases will continue to decrease. Asymptomatic infections arising in the next four months also showed a similar trend compared to symptomatic infections across different VR and vaccination effectiveness, albeit at a higher magnitude (Fig S3). Different vaccination rates would also alter the trajectory of cumulative COVID-19 related death, where more deaths could be prevented given a higher VR (Fig S4).

**Figure 2.**
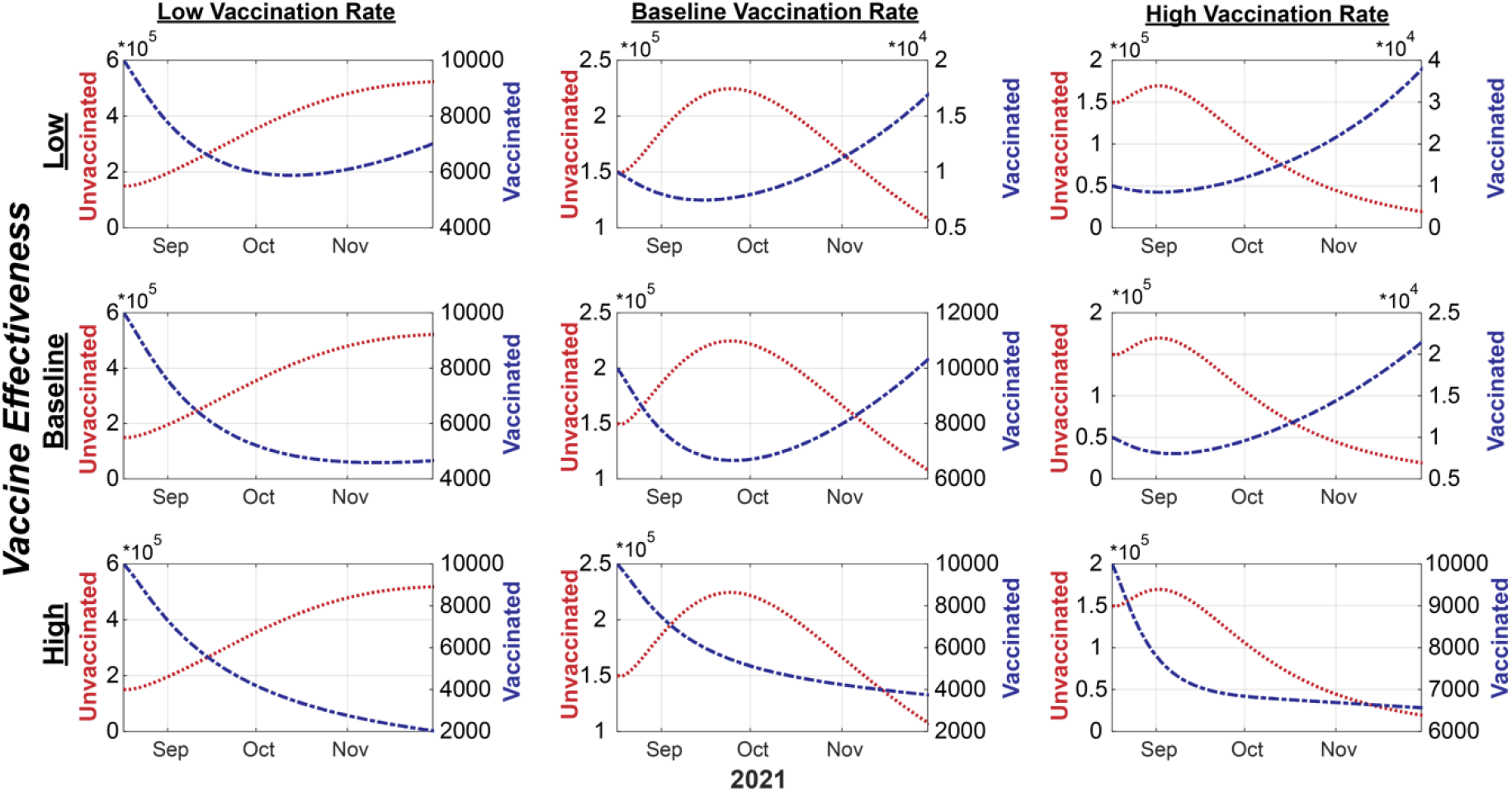
Modelled projections of symptomatic infections in unvaccinated and vaccinated subpopulations under different vaccination rates and vaccine effectiveness. The case counts of the unvaccinated individuals are depicted by the red line and the unit labels on the left-side y-axis, whereas the infected, vaccinated (breakthrough) cases are depicted by the blue line and the unit labels on the right-side y-axis.

### The control reproduction number mediated by natural and vaccine-induced immunity

A pandemic decline when the control reproduction number (*R*_*c*_)—the average number of new infections generated by an infected individual over the infected period in a controlled population (i.e., one with a vaccination programs)—is lower than one (*R*_*c*_ < 1). The disease-free equilibrium may be achieved when sufficient fractions of the populations are fully vaccinated and recovered (natural immunity) individuals. We derived *R*_*c*_ and obtained its value under different levels of transmission rates and vaccine effectiveness (supplementary methods). Under a baseline transmission rate and given that 30% of the population recovered from the disease and acquired natural immunity *(10)*, over 54%, 41% and 46% of the simulated US population would need to be fully vaccinated to achieve *R*_*c*_ < 1 given low, baseline, and high vaccine effectiveness, respectively (Fig 3). The required fractions of fully vaccinated individuals to reduce the value of *R*_*c*_ lower than one are 44%, 42%, and 38% under low levels of transmission, and increase to near-saturated 66%, 62%, and 56% under high levels of transmission. We also obtained the *R*_*c*_ for asymptomatic infections, which generally showed higher requirements of vaccinated individuals due to the lower vaccine effectiveness against asymptomatic infections (Fig S6).

**Figure 2.**
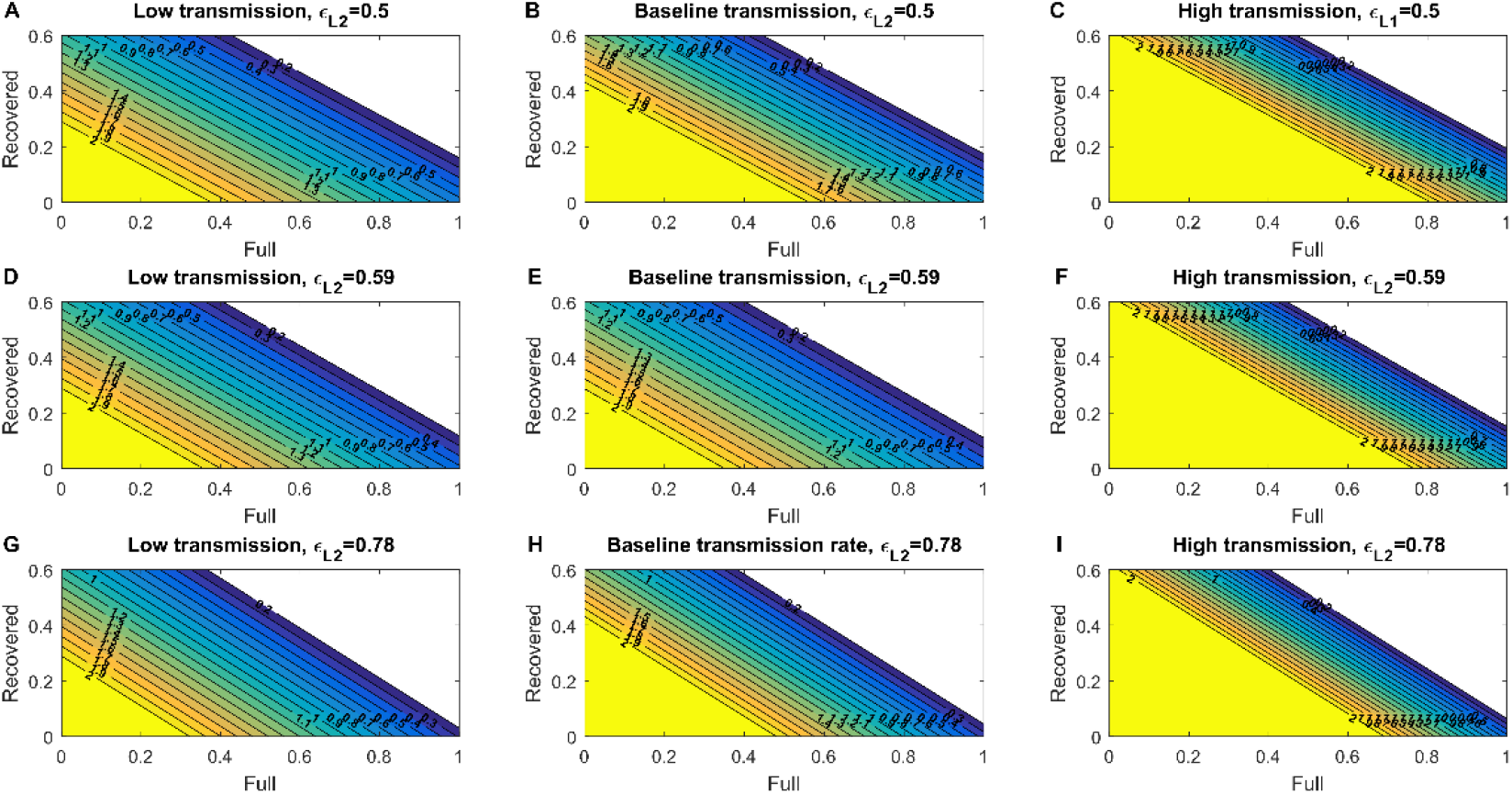
Control Reproduction number for symptomatic transmission considering full immunity and recovered individuals. The assumed vaccine effectiveness (ε_L2_) parameter under each scenario is denoted on top of each panel and are inferred based on real-world data of individuals receiving two doses of the BNT162b2 vaccine (9). The heatmaps in left row consider a low transmission rate; the heatmaps in the center row panels consider a baseline transmission rate; and the heatmaps in the right row panels consider a high transmission rate.

To model another scenario where the vaccine effectiveness reduces further due to factors such as a more resistant virus variant, we evaluated *R*_*c*_ using the vaccine effectiveness parameters associated with one dose of the vaccine. The disease-free equilibrium in this population is composed of susceptible, recovered and partially immune (equivalent to one dose vaccine’s immunity) individuals. Under a baseline transmission rate and given that 30% of the population recovered from the disease and acquired natural immunity, over 82%, 56%, and 67% of the simulated US population would need to have vaccine-induced, partial immunity to achieve *R*_*c*_ < 1 given low, baseline, and high vaccine effectiveness, respectively (Fig 4). The required fractions of vaccinated individuals reduce to 68%, 58%, and 55% under low levels of transmission, and increase to near-saturated levels of 100%, 86%, and 82% under high levels of transmission (Fig 4). Thus, at this level of vaccine effectiveness, a combination of measures to lower virus transmission would be required in conjunction with natural and vaccine-induced immunity to diminish the pandemic.

**Figure 4.**
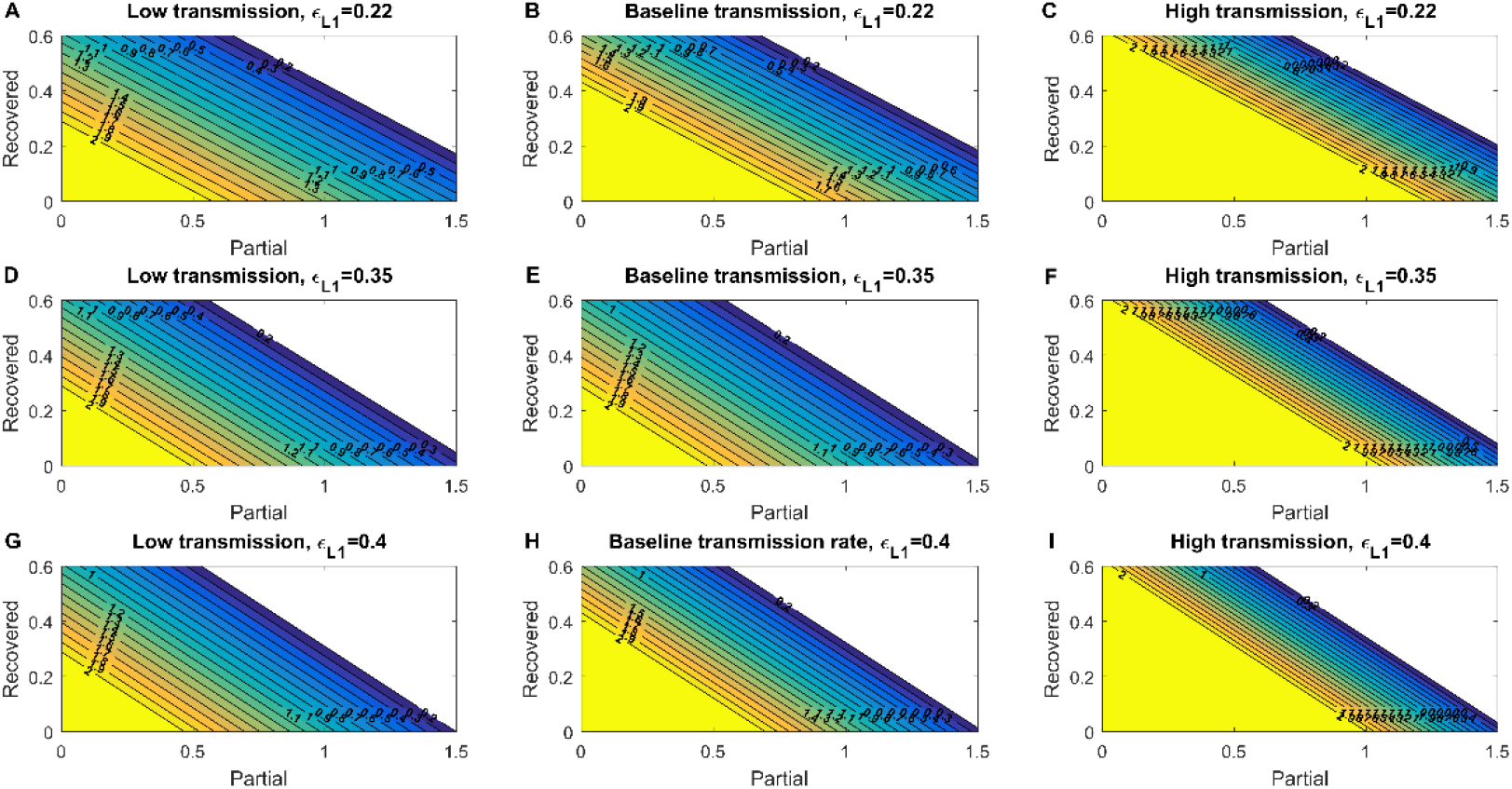
Control Reproduction number for symptomatic transmission considering partial immunity and recovered individuals. The assumed vaccine effectiveness (ε_L1_) parameter under each scenario is denoted on top of each panel and are inferred based on real-world data of individuals acquiring partial immunity as those induced by one does of the BNT162b2 vaccine (9). The heatmaps in left row consider a low transmission rate; the heatmaps in the center row panels consider a baseline transmission rate; and the heatmaps in the right row panels consider a high transmission rate.

### Sensitivity analyses of model parameters affecting the pandemic trajectories

To identify the transmission and vaccination factors that may affect the spread of the delta variant, we conducted sensitivity analyses *(11)* to determine how changes in each modelled parameter may alter the output *R*_*c*_. In the local sensitivity analysis, the elastic index of each parameter was obtained by applying its partial derivatives and substitution of its value one at a time (material and methods). As expected, increasing the force of infection for symptomatic (*β*_1_) and asymptomatic (*β*_2_) infections, along with the proportions of symptomatic individuals in unvaccinated and vaccine infections (*p*_1_, *p*_2_), are implicated in increasing the *R*_*c*_ related to symptomatic infections. On the other hand, a higher recuperation rate (*γ*) could significantly reduce *R*_*c*_, likely due to the natural immunity acquired by infected individuals. Increases in vaccine effectiveness parameters for both one or two doses, as well as increasing vaccination rate (*ρ*), would lower the *R*_*c*_ and help control of the pandemic (Fig 5).

**Figure 3.**
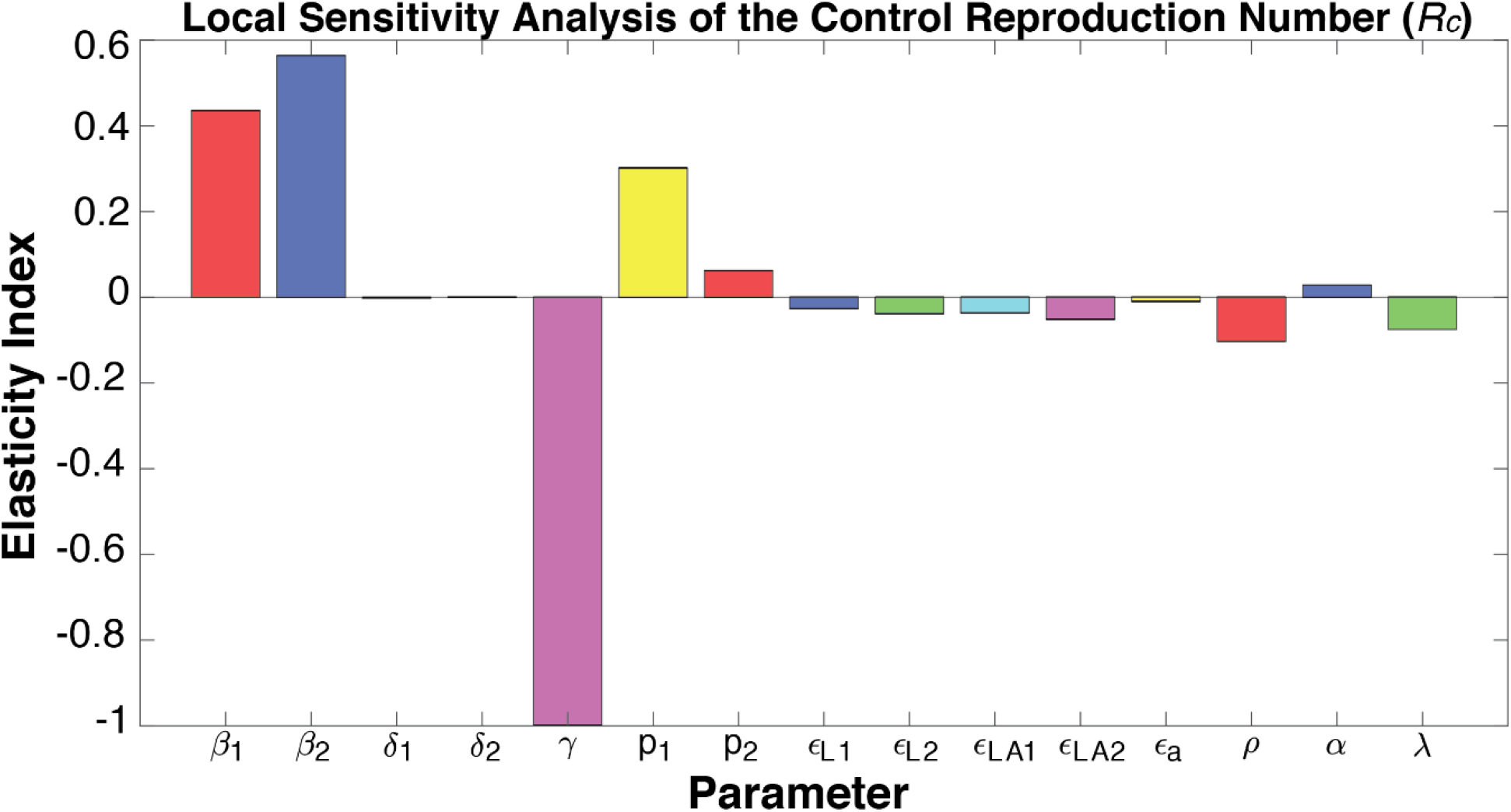
Elasticity index showing each modelled parameter’s effect on the control reproduction number as determined in the local sensitivity analysis. For each column, a positive value represents an increase of the parameter is associated with an increment in the control reproduction number, whereas a negative value means a decrease.

While local sensitivity analysis can help determine each factor’s impact one at a time, input parameters can show drastic changes or spontaneously shape model behavior and outputs based on their interactions. To model their concerted impact on the number of symptomatic infections in both vaccinated and unvaccinated individuals, we used a global sensitivity approach that computes the partial rank correlation coefficient (PRCC) by sampling with the Latin hypercube method *(12)*. For symptomatic infections in the unvaccinated individuals, the force of infection (*β*_1_, *β*_2_) is positively correlated with increasing the dynamics for this population (Fig 6A) and it is statically significant (Fig S6A). The recuperation rate (γ) is negatively correlated with newly infected cases, likely due to the natural immunity developed in recovered individuals. As the analyses focused on symptomatic individuals, *p*_1_ by definition, shows a significant positive correlation (Fig 6A and Fig S6A). Model parameters that determine asymptomatic infections in unvaccinated individuals behave in a similar manner as symptomatic infections (Fig S7A and B). We also conducted global sensitivity analyses to identify how parameter changes may affect the sub-populations with partial or full vaccine-induced immunity. Partial immunity individuals are influenced negatively by the force of infection for symptomatic and asymptomatic infections as well as *p*_1_ indicating the fraction of symptomatic infections, while the recuperation rate is positively correlated with partial immunity individuals (Fig S8A and B). Full immunity individuals, derived from partial-immunity individuals who proceed to receive the second vaccination shot, are influenced in a similar manner (Fig S9A and B).

**Figure 4.**
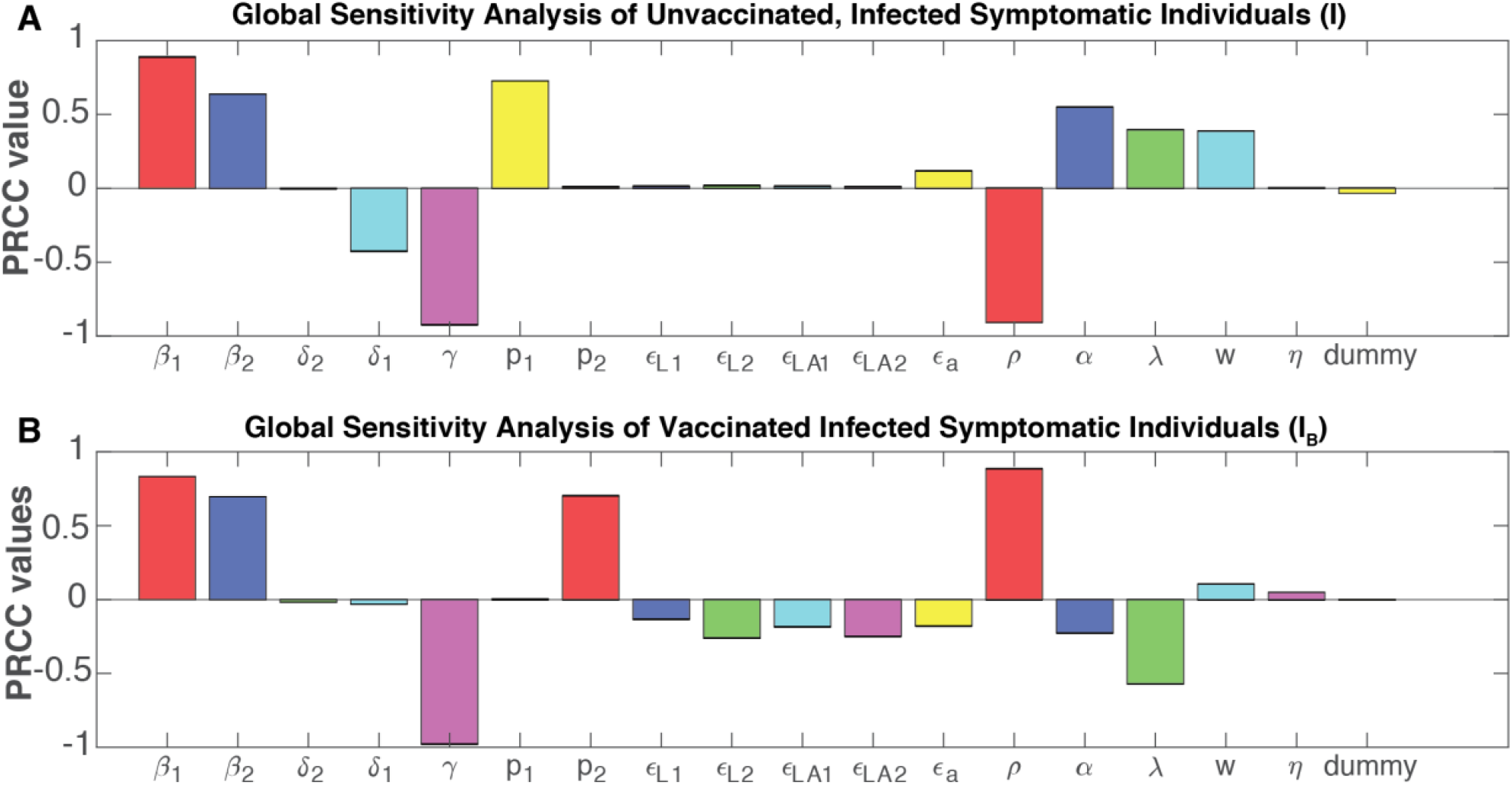
Global sensitivity analyses of the parameters correlated with the symptomatic infections in (A) unvaccinated, and (B) vaccinated individuals.The relationship between each parameter and the infected subpopulation is measured by the Partial Rank Correlation Coefficient (PRCC); a positive PRCC indicates an increase in the parameters is correlated with an increase of the said sub-population, and a negative PRCC vice versa. The significance level of each PRCC is indicated in Fig. S6.

As increasing fractions of the global population become vaccinated, we further conducted global sensitivity analyses to identify parameters associated with diminishing symptomatic infections in vaccinated individuals (i.e., breakthrough cases). Similar to symptomatic infections in unvaccinated individuals, breakthrough cases are positively correlated with the forces of infection for either symptomatic or asymptomatic, and negatively correlated with recuperation rate (γ) (Fig 6B). As expected, each of the vaccine effectiveness parameters is negatively correlated with breakthrough cases. A higher vaccination rate is correlated with increased breakthrough cases given it results in a larger pool of vaccinated individuals, but it also shows a strong negative correlation with infections in unvaccinated individuals, which can be ∼10-fold higher (Fig 2). On the other hand, a high waning rate (*α*) for vaccine-induced immunity is correlated with a sizable increase in infections in unvaccinated individuals but reduced infections in vaccinated individuals, likely due to the model’s turn-over of vaccinated individuals back to the susceptible individuals upon complete immune waning.

## Discussion

Mathematical models can forecast the spread of infectious diseases and help inform public health decisions. Here, we develop a unique compartmentalized model that considers virus spread in unvaccinated and vaccinated subpopulations to better portray the ongoing COVID-19 pandemic. Our timely application of the model to the US population considers transmission and immunity parameters associated with the currently-dominant delta variant, a two-dose vaccination scheme, and the variant’s partial resistance to the vaccine that contributed to breakthrough cases.

Over half of the US population have been fully vaccinated in September 2021. Despite the larger fraction of the vaccinated compared to unvaccinated individuals, the model showed that the number of infected individuals would be a magnitude higher (∼10x) in the unvaccinated compared to the vaccinated subpopulation within the ranges of likely transmission rates and vaccine effectiveness. Vaccinating a larger fraction of the population, in conjunction with practicing NPIs, will continue to be one of the most effective means to diminish SARS-CoV-2 transmissions (*13*). Vaccine-induced immunity provided by two doses is more effective compared to one dose to reduce disease transmission or COVID-19 related hospitalizations/deaths (*14*). Meanwhile, the reduction in vaccine effectiveness against virus variants and waning immunity may require additional solutions. We note that the vaccine waning parameter used herein is approximated, and while there are evidence supporting the loss of protection, the compounding effects of a more infectious variant (i.e., delta) and waning immunity can be difficult to dissect *(15)*. Nevertheless, rapid development of new vaccines or and administration a third, bolster dose *(16)* are active areas of research that may yield promising results for controlling the pandemic.

The development of the model herein is informed by multiple parameters of virus transmission and vaccine-induced immunity, which have resulted from consistent monitoring and rapid sharing of data throughout the COVID-19 pandemic. For example, the model’s case counts and vaccination rates were established using repository data of daily new COVID-19 infections and daily vaccination doses applied in the US *(8)*. The parameters of the BNT162b2 vaccine’s immunity against the delta variant (either one or two doses) were obtained from the real-world data (*9*) determined in UK, which when applied to the US pandemic, allowed us to circumvent the potential circular logic of forecasting the pandemic using vaccine effectiveness parameters derived from the same population. Scientific progress in the ongoing COVID-19 pandemic is accelerated by the promptness and transparency in data sharing practices that could also help tackle a wide array of public health challenges.

Our model has several limitations, some of which are addressed in other mathematical models developed during this pandemic. First, our model does not consider the population heterogeneity, which has been shown to reduce the required number of vaccinated individuals to achieve herd immunity (*7*). Second, we do not consider seasonality (*6*), although the seasonal trend for SARS-CoV-2 is still unclear at this point. Third, SARS-CoV-2 infections that are not reported were not included in the model and may vary based on ascertainment rates *(17)*. Lastly, we focused the application on modeling the currently dominant delta variant in the US as of September 2021. While the analyses herein utilized the immunity parameters for the BNT162b2 vaccine and the delta variant, our model can be applied to model other vaccines and emerging virus strains in different populations.

Overall, our results demonstrated that the trajectory of a pandemic is heavily influenced by natural and vaccine-induced immunity given a dominant virus strain’s level of infectiousness and response to vaccines. As variants of SARS-CoV-2 have emerged (*18*), VOCs capable of higher transmission rates and immune evasion may continue to arise (*19-21*). Tempering the spread of SARS-CoV-2 variants will require enhanced global efforts on sequencing and variant detection, establishing reproducible analysis pipelines, and rapid sharing of data across geographical boundaries (*22*). Given that asymptomatic but infected virus carriers can spread SARS-CoV-2 (*23,24*), increased testing and surveillance will also be critical. Finally, to complement the partial protection provided by vaccination programs, practicing NPIs including social distancing, imitating indoor group gatherings, and wearing face masks (25) can help reduce transmission rates. The emergence of new SARS-CoV-2 strains has introduced unique challenges in the ongoing pandemic, but may also spur innovations that can help control infectious virus variants in the future.

## Data Availability

The original contributions presented in the study are included in the article/Supplementary Material. All code used for analysis is available https://github.com/UgoAvila/Delta-Variant-In-the-US.

## Acknowledgments

Ugo Avila Ponce de León is a doctoral student from Programa de Doctorado en Ciencias Biológicas of the Universidad Nacional Autónoma de México (UNAM). This paper was developed in the period of his Ph.D. studies.

## Author contributions

**UAPL:** Conceptualization, Methodology, Software, Formal analysis, Data curation, Writing - original draft, Writing - review & editing. **EAV:** Conceptualization, Methodology, Supervision, Project administration. **KLH:** Conceptualization, Methodology, Writing-review & editing. Project administration.

## Data and materials availability

Supplementary Materials

Materials and Methods

Supplementary Text

Figs. S1 to S9

Tables S1

## References

1. E. Dong, H. Du, L. Gardner, An interactive web-based dashboard to track COVID-19 in real time. Lancet Infect. Dis. 20, 533–534 (2020).

2. A. S. Lauring, E. B. Hodcroft, Genetic Variants of SARS-CoV-2—What Do They Mean? JAMA. 325, 529–531 (2021).

3. S. Elbe, G. Buckland-Merrett, Data, disease and diplomacy: GISAID’s innovative contribution to global health. Glob Chall. 1, 33–46 (2017).

4. Eurosurveillance editorial team, Updated rapid risk assessment from ECDC on the risk related to the spread of new SARS-CoV-2 variants of concern in the EU/EEA -first update. Euro Surveill. 26 (2021), doi:10.2807/1560-7917.ES.2021.26.3.2101211.

5. E. Callaway, Delta coronavirus variant: scientists brace for impact. Nature. 595, 17–18 (2021).

6. C. M. Saad-Roy, C. E. Wagner, R. E. Baker, S. E. Morris, J. Farrar, A. L. Graham, S. A. Levin, M. J. Mina, C. J. E. Metcalf, B. T. Grenfell, Immune life history, vaccination, and the dynamics of SARS-CoV-2 over the next 5 years. Science. 370, 811–818 (2020).

7. T. Britton, F. Ball, P. Trapman, A mathematical model reveals the influence of population heterogeneity on herd immunity to SARS-CoV-2. Science. 369, 846–849 (2020).

8. E. Mathieu, H. Ritchie, E. Ortiz-Ospina, M. Roser, J. Hasell, C. Appel, C. Giattino, L. Rodés-Guirao, A global database of COVID-19 vaccinations. Nat Hum Behav. 5, 947–953 (2021).

9. P. Elliott, D. Haw, H. Wang, O. Eales, C. Walters, K. Ainslie, C. Atchison, C. Fronterre, P. Diggle, A. Page, Others, REACT-1 round 13 final report: exponential growth, high prevalence of SARS-CoV-2 and vaccine effectiveness associated with Delta variant in England during May to July 2021 (2021) (available at https://spiral.imperial.ac.uk/handle/10044/1/90800).

10. S. Pei, T. K. Yamana, S. Kandula, M. Galanti, J. Shaman, Burden and characteristics of COVID-19 in the United States during 2020. Nature (2021), doi:10.1038/s41586-021-03914-4.

11. H. S. Rodrigues, M. T. T. Monteiro, D. F. M. Torres, Sensitivity analysis in a dengue epidemiological model. Conf. Pap. Math. 2013, 1–7 (2013).

12. S. Marino, I. B. Hogue, C. J. Ray, D. E. Kirschner, A methodology for performing global uncertainty and sensitivity analysis in systems biology. J. Theor. Biol. 254, 178–196 (2008).

13. L. Matrajt, H. Janes, J. T. Schiffer, D. Dimitrov, Quantifying the Impact of Lifting Community Nonpharmaceutical Interventions for COVID-19 During Vaccination Rollout in the United States. Open Forum Infect Dis. 8, ofab341 (2021).

14. L. Matrajt, J. Eaton, T. Leung, D. Dimitrov, J. T. Schiffer, D. A. Swan, H. Janes, Optimizing vaccine allocation for COVID-19 vaccines shows the potential role of single-dose vaccination. Nat. Commun. 12, 3449 (2021).

15. H. L. Moline, M. Whitaker, L. Deng, J. C. Rhodes, J. Milucky, H. Pham, K. Patel, O. Anglin, A. Reingold, S. J. Chai, N. B. Alden, B. Kawasaki, J. Meek, K. Yousey-Hindes, E. J. Anderson, M. M. Farley, P. A. Ryan, S. Kim, V. T. Nunez, K. Como-Sabetti, R. Lynfield, D. M. Sosin, C. McMullen, A. Muse, G. Barney, N. M. Bennett, S. Bushey, J. Shiltz, M. Sutton, N. Abdullah, H. K. Talbot, W. Schaffner, R. Chatelain, J. Ortega, B. P. Murthy, E. Zell, S. J. Schrag, C. Taylor, N. Shang, J. R. Verani, F. P. Havers, Effectiveness of COVID-19 Vaccines in Preventing Hospitalization Among Adults Aged ≥65 Years - COVID-NET, 13 States, February-April 2021. MMWR Morb. Mortal. Wkly. Rep. 70, 1088–1093 (2021).

16. [No title], (available at https://www.fda.gov/media/144413/download).

17. W. T. Harvey, A. M. Carabelli, B. Jackson, R. K. Gupta, E. C. Thomson, E. M. Harrison, C. Ludden, R. Reeve, A. Rambaut, S. J. Peacock, D. L. Robertson, COVID-19 Genomics UK (COG-UK) Consortium, SARS-CoV-2 variants, spike mutations and immune escape. Nature Reviews Microbiology. 19 (2021), pp. 409–424.

18. T. Tada, H. Zhou, B. M. Dcosta, M. I. Samanovic, M. J. Mulligan, N. R. Landau, SARS-CoV-2 Lambda Variant Remains Susceptible to Neutralization by mRNA Vaccine-elicited Antibodies and Convalescent Serum. bioRxiv (2021), p. 2021.07.02.450959.

19. I. Kimura, Y. Kosugi, J. Wu, D. Yamasoba, E. P. Butlertanaka, Y. L. Tanaka, Y. Liu, K. Shirakawa, Y. Kazuma, R. Nomura, Y. Horisawa, K. Tokunaga, A. Takaori-Kondo, H. Arase, The Genotype to Phenotype Japan (G2P-Japan) Consortium, A. Saito, S. Nakagawa, K. Sato, SARS-CoV-2 Lambda variant exhibits higher infectivity and immune resistance. bioRxiv (2021), p. 2021.07.28.454085.

20. M. McCallum, J. Bassi, A. De Marco, A. Chen, A. C. Walls, J. Di Iulio, M. A. Tortorici, M.-J. Navarro, C. Silacci-Fregni, C. Saliba, K. R. Sprouse, M. Agostini, D. Pinto, K. Culap, S. Bianchi, S. Jaconi, E. Cameroni, J. E. Bowen, S. W. Tilles, M. S. Pizzuto, S. B. Guastalla, G. Bona, A. F. Pellanda, C. Garzoni, W. C. Van Voorhis, L. E. Rosen, G. Snell Telenti, H. W. Virgin, L. Piccoli, D. Corti, D. Veesler, SARS-CoV-2 immune evasion by the B.1.427/B.1.429 variant of concern. Science. 373, 648–654 (2021).

21. N. D. Grubaugh, E. B. Hodcroft, J. R. Fauver, A. L. Phelan, M. Cevik, Public health actions to control new SARS-CoV-2 variants. Cell. 184, 1127–1132 (2021).

22. C. P. Muller, Do asymptomatic carriers of SARS-COV-2 transmit the virus? Lancet Reg Health Eur. 4, 100082 (2021).

23. P. Wilmes, J. Zimmer, J. Schulz, F. Glod, L. Veiber, L. Mombaerts, B. Rodrigues, A. Aalto, J. Pastore, C. J. Snoeck, M. Ollert, G. Fagherazzi, J. Mossong, J. Goncalves, A. Skupin, U. Nehrbass, SARS-CoV-2 transmission risk from asymptomatic carriers: Results from a mass screening programme in Luxembourg. Lancet Reg Health Eur. 4, 100056 (2021).

24. Y. Cheng, N. Ma, C. Witt, S. Rapp, P. S. Wild, M. O. Andreae, U. Pöschl, H. Su, Face masks effectively limit the probability of SARS-CoV-2 transmission. Science (2021), doi:10.1126/science.abg6296.

