## Supplementary Materials for "Modeling the transmission of the SARS-CoV-2 delta variant in a partially vaccinated population"

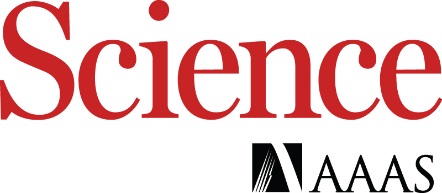

Supplementary Materials for

Modeling the transmission of the SARS-CoV-2 delta variant in a partially vaccinated population

Ugo Avila-Ponce de León, Eric Avila-Vales, Kuan-lin Huang

**This PDF file includes:**

Materials and Methods

Supplementary Text

Figs. S1 to S6

Table S1

Materials and Methods

**3.1 Parameter Estimation for the Mathematical Model**

To test the utility of this delta variant two doses vaccine model, we tested its application on simulating the dynamics of new infections of COVID-19 in the US. We will assume that the infection, recovery rate and death rate are time dependent functions like in (*1*, *2*). For the estimation of the parameters we use the cumulative data provided by the Johns Hopkins University repository (*3*).  For estimating parameters for the delta variant we considered the data of COVID-19 infections, recovery and death from the period of June 2021 to 17^th^ August 2021, period where delta variant was the dominant strain throughout the US (*4*). Parameter estimation for the delta variant was pursued by applying a vaccination rate equal to zero and the subpopulation associated with the alpha variant equal to cero. For this approach there were only two fixed parameters the average length of the latent period and the proportion of symptomatic individuals. The system of differential equations was solved using Matlab R2016b and ODE45 solver. The parameters were fitted by applying two methods, i.e., the ones described in (*1*, *2*) and (*5*). The code and implementation can be downloaded at <https://github.com/UgoAvila/Delta-Variant-In-the-US>

**3.2. Local Sensitivity Analysis of the Parameters of the Mathematical Model**

The sensitivity analysis of the control reproduction number was carried out using the following definition (*6*): If $R_{c}$ is differentiable with respect to a given parameter $\theta$, then the normalized forward sensitivity index of $R_{c}$ is defined by:

$$\Gamma_{\theta}^{R_{c}}=\frac{\theta}{R_{c}}*\frac{\partial R_{c}}{\partial\theta}.$$

Once we obtained the parameters, we were interested to perturb them to determine how their changes affect the reproduction number, we solved the equations using Mathematica. After we had the partial derivation of a parameter, we fixed the value of the other parameters fitted from data or obtained from the clinical trials and computed the index of the parameters. A positive sign of the parameter correlates with an increase of the control reproduction, whereas a negative sign is associated with a decrease of the control reproduction number.

**3.3. Global Sensitivity Analysis of the Mathematical Model**

In this subsection, we applied and adapted the global sensitivity analysis approach described in (*7*) to our model*.* We sampled the 17 included parameters and evaluated which were important in determining the behavior of our model. We applied the analysis by using partial rank correlation coefficient (PRCC) by sampling using the Latin hypercube method. These methods allow us to evaluate which parameter affects our response function. In our case, we used the eleven differential equations as our response function. The PRCC values range between -1 and 1, where a negative value means that parameter is negatively correlated with the response function evaluated, and a positive value suggests a positive correlation between the parameter with the response function be evaluated. The significance of each correlation is evaluated, and we only report parameters whose correlations with the response function show a p-value $\leq0.05$.

Supplementary Text

**Mathematical Model with Asymptomatic individuals and Vaccination.**

We develop and apply a compartmental differential equation to understand the dynamics of the spread of the delta variant in vaccinated and non-vaccinated individuals. The mathematical model contains two separate sets of equations for the virus strains that affect the same susceptible population and vaccinated subpopulation. Our SPFEIARD model evaluates the dynamics of ten subpopulations at any given time $t$, which are denoted as: $S\left( t \right), E_{1}(t)$, etc.  Fig 1 shows a diagram of the flow through the compartmentalized subpopulations.

**Susceptible population S(t)**: Because we are not considering any natural recruitments (births), this subpopulation cannot increase in this manner. Susceptible population vaccinated or not become exposed with a symptomatic delta variant infection at a rate $\beta_{1}$ and an asymptomatic delta variant infection at a rate $\beta_{2}$. The susceptible population may increase at a rate $\alpha$ associated with the waning rate of a vaccine, a parameter of an imperfect vaccine. Finally, the susceptible population will be vaccinated at a rate $\rho\geq0$, multiplied by the all or nothing protection of a vaccine ${(1-\varepsilon}_{a})$. The rate of change of the susceptible population is expressed in the following equation:

$$\frac{dS}{dt}=-\beta_{1}\left( \frac{I+I_{B}}{N} \right)S-\beta_{2}\left( \frac{A+A_{B}}{N} \right)S+\alpha F+\eta R-\left( 1-\varepsilon_{a} \right)\rho S. (S1)$$

**Population exposed to the original variant,** $\boldsymbol{E(t)}$: Because we are simulating an infection caused by a virus, we need to include the exposed population. This subpopulation are individuals that are infected but not infectious. It increases at a rate of infection $\beta_{1}$ and $\beta_{2}$ for symptomatic and asymptomatic infection from the delta variant. This subpopulation evaluated the dynamics of new infections of individuals that have not receive any type of vaccine. This population decreases at a rate $w$, which denotes the average length of the latent period. Hence,

$$\frac{dE}{dt}=\left( \frac{\beta_{1}I}{N} \right)S+\left( \frac{\beta_{2}A}{N} \right)S-wE. (S2)$$

**Population infected by the original variant,** $\boldsymbol{I(t)}$: Infected individuals that develop symptoms for the delta variant are generated at a proportion $p_{1}\in(0,1)$ from the exposed non-vaccinated subpopulation. They recover at a rate $\gamma$ and die from the disease at a rate $\delta_{1}$. Consequently,

$$\frac{dI}{dt}=p_{1}wE-\left( \delta_{1}+\gamma\right)I. (S3)$$

**Asymptomatic population infected by the original variant,** $\boldsymbol{A(t)}$: This population is considered an infected population but do not develop symptoms of COVID-19. They are important to include in our model because they can spread the virus, they are generated at a proportion $(1-p_{1})$ from the exposed class. This population recovers at a rate$\gamma$. This population does not die from this disease because they do develop symptoms. So,

$$\frac{dA}{dt}=(1-p_{1})wE-\gamma A. (S4)$$

**Population of individuals with partial immunity,** $\boldsymbol{P(t)}$**:** This subpopulation is vaccinated at a rate $\rho\leq0$ multiplied by the all or nothing protection $\varepsilon_{a}$. This population decreases at a rate $\left( 1-\varepsilon_{L1} \right)\beta_{1}$ due to leakiness or vaccine efficiency provided by one dose of the Pfizer vaccine for symptomatic individuals. The vaccine efficiency protections extend to the asymptomatic infected individuals as well, so it decays at a rate $\left( 1-\varepsilon_{LA1} \right)\beta_{2}$. Finally, this population decreases when individuals receive the second dose of the Pfizer vaccine at a rate $\lambda$. Hence,

$$\frac{dP}{dt}=\left( 1-\varepsilon_{a} \right)\rho S-\left( \frac{\left( 1-\varepsilon_{L1} \right)\beta_{1}\left( I+I_{B} \right)}{N} \right)P-\left( \frac{\left( 1-\varepsilon_{LA1} \right)\beta_{2}\left( A+A_{B} \right)}{N} \right)P-\lambda P. (S5)$$

**Population of individuals with full immunity,** $\boldsymbol{F(t)}$**:** This population increases at a rate $\lambda$ but for it to increases it is mandatory that individuals first they are part of the partial immunity subpopulation. Due to the imperfect of the vaccine, this population decays at a rate $\left( 1-\varepsilon_{L2} \right)\beta_{1}$ for symptomatic infected individuals due to the interaction with individuals infected with the delta variant. As well it decays at a rate $\left( 1-\varepsilon_{LA2} \right)\beta_{2}$ for asymptomatic infected individuals due to the interaction with individuals infected with the delta variant. Finally, the population decays as well at a rate $\alpha$ due to the loss of protection or waning immunity provided by the protection of the vaccine.

$$\frac{dF}{dt}=\lambda P-\left( \frac{\left( 1-\varepsilon_{L2} \right)\beta_{1}\left( I+I_{B} \right)}{N} \right)F-\left( \frac{\left( 1-\varepsilon_{LA2} \right)\beta_{2}\left( A+A_{B} \right)}{N} \right)F-\alpha F. (S6)$$

**Population exposed to the delta variant while being vaccinated with one or two doses,** $\boldsymbol{E}_{\boldsymbol{B}}\boldsymbol{(t)}$: We include the exposed vaccinated population to the delta variant because we are modeling a virus and we need to include the latent period. The population enlarges at a rate $\beta_{1}$ for symptomatic infected individual and at a rate $\beta_{2}$ for asymptomatic individuals, both from the delta variant. Moreover, vaccinated individuals who interact with symptomatic or asymptomatic are incorporated at a rate $\left( 1-\varepsilon_{L1} \right)$ and $\left( 1-\varepsilon_{LA1} \right)$ respectively, which is the vaccine efficiencies of the vaccine to the delta variant for one dose. Also, vaccinated individuals who interact with symptomatic or asymptomatic are incorporated at a rate $\left( 1-\varepsilon_{L2} \right)$ and $\left( 1-\varepsilon_{LA2} \right)$ respectively which is the vaccine efficiencies of the vaccine to the delta variant for two dose. This subpopulation deceases at a rate $w$, the length were individuals pass from infected to infected and infectious. Hence,

$$\frac{dE_{B}}{dt}=\left( \frac{\left( 1-\varepsilon_{L1} \right)\beta_{1}\left( I+I_{B} \right)}{N} \right)P+\left( \frac{\left( 1-\varepsilon_{LA1} \right)\beta_{2}\left( A+A_{B} \right)}{N} \right)P+\left( \frac{\left( 1-\varepsilon_{L2} \right)\beta_{1}\left( I+I_{B} \right)}{N} \right)F+\left( \frac{\left( 1-\varepsilon_{LA2} \right)\beta_{2}\left( A+A_{B} \right)}{N} \right)F-wE_{B}. (S7)$$

**Population infected by the delta variant while being vaccinated,** $\boldsymbol{I}_{\boldsymbol{B}}\boldsymbol{(t)}$: All individuals that are infected while being vaccinated regardless of the dose received and develop symptoms at a proportion$p_{2}\in\left( 0,1 \right)$ who will recover at a rate $\gamma$ and die at a rate $\delta_{2}$. Consequently,

$$\frac{dI_{B}}{dt}=p_{2}wE_{B}-\left( \delta_{2}+\gamma\right)I_{B} . (S8)$$

**Asymptomatic population infected by the delta variant while being vaccinated,** $\boldsymbol{A}_{\boldsymbol{B}}\boldsymbol{(t)}$: individuals that are infected while being vaccinated regardless of the dose received but do not develop symptoms at a proportion $(1-p_{2})$ from the exposed class. They recover at a rate$\gamma$. So,

$$\frac{dA_{B}}{dt}=\left( 1-p_{2} \right)wE_{B}-\gamma A_{B}. (S9)$$

**Recovered population,** $\boldsymbol{R(t)}$: All individuals infected vaccinated or not with symptoms or not will recover at a rate $\gamma$ from the delta variant. We acknowledge in this population the loss of protection from natural immunity at a rate $\eta$. Consequently,

$$\frac{dR}{dt}=\gamma\left( {I+A+I}_{B}+A_{B} \right)-\eta R. (S10)$$

**Death population,** $\boldsymbol{D(t)}$: Infected individuals with symptoms die at a rate $\delta_{1}$ from the delta variant and $\delta_{2}$ are vaccinated individuals that die due to the delta variant. So,

$$\frac{dD}{dt}=\delta_{1}I_{1}+\delta_{2}I_{B} . (S11)$$

Henceforth, the system of differential equations that will simulate the dynamics of the original strain and the alpha variant with vaccination is:

$$\frac{dS}{dt}=-\beta_{1}\left( \frac{I+I_{B}}{N} \right)S-\beta_{2}\left( \frac{A+A_{B}}{N} \right)S+\alpha F+\eta R-\left( 1-\varepsilon_{a} \right)\rho S,$$

$$\frac{dE}{dt}=\left( \frac{\beta_{1}I}{N} \right)S+\left( \frac{\beta_{1}A}{N} \right)S-wE,$$

$$\frac{dI}{dt}=p_{1}wE-\left( \delta_{1}+\gamma\right)I,$$

$$\frac{dA}{dt}=(1-p_{1})wE-\gamma A, (S12)$$

$$\frac{dP}{dt}=\left( 1-\varepsilon_{a} \right)\rho S-\left( \frac{\left( 1-\varepsilon_{L1} \right)\beta_{1}\left( I+I_{B} \right)}{N} \right)P-\left( \frac{\left( 1-\varepsilon_{LA1} \right)\beta_{2}\left( A+A_{B} \right)}{N} \right)P-\lambda P,$$

$$\frac{dF}{dt}=\lambda P-\left( \frac{\left( 1-\varepsilon_{L2} \right)\beta_{1}\left( I+I_{B} \right)}{N} \right)F-\left( \frac{\left( 1-\varepsilon_{LA2} \right)\beta_{2}\left( A+A_{B} \right)}{N} \right)F-\alpha F,$$

$$\frac{dE_{B}}{dt}=\left( \frac{\left( 1-\varepsilon_{L1} \right)\beta_{1}\left( I+I_{B} \right)}{N} \right)P+\left( \frac{\left( 1-\varepsilon_{LA1} \right)\beta_{2}\left( A+A_{B} \right)}{N} \right)P+\left( \frac{\left( 1-\varepsilon_{L2} \right)\beta_{1}\left( I+I_{B} \right)}{N} \right)F+\left( \frac{\left( 1-\varepsilon_{LA2} \right)\beta_{2}\left( A+A_{B} \right)}{N} \right)F-wE_{B},$$

$$\frac{dI_{B}}{dt}=p_{2}wE_{B}-\left( \delta_{2}+\gamma\right)I_{B},$$

$$\frac{dA_{B}}{dt}=\left( 1-p_{2} \right)wE_{B}-\gamma A_{B},$$

$$\frac{dR}{dt}=\gamma\left( {I+A+I}_{B}+A_{B} \right)-\eta R,$$

$$\frac{dD}{dt}=\delta_{1}I_{1}+\delta_{2}I_{B}.$$

We can derive that $N(t)$ at a time $t$ is given by,

$$N\left( t \right)=S\left( t \right)+E\left( t \right)+I\left( t \right)+A\left( t \right)+P\left( t \right)+F(t)+E_{B}\left( t \right)+I_{B}\left( t \right)+A_{B}\left( t \right)+R\left( t \right)+D\left( t \right)$$

**2.1 Control reproduction number with a disease-free equilibrium**

In our mathematical model, there exists a disease-free equilibrium which happens when the vaccination rate $\rho\neq0.$ So $E=I=A=E_{B}=I_{B}=A_{B}=0$, and the disease-free equilibrium is:

$$x_{DF}=\left( S^{*},0,0,0,P^{*},F^{*},0,0,0,0,0 \right).$$

Where $S^{*}\geq0$ , $P^{*}>0$ and $F^{*}>0$.

This equilibrium represents where the vaccination logistic program has ended, with a certain number of individuals that are vaccinated, and a population that may be susceptible to get infected. We will compute the control reproduction number $R_{c}$ in this disease-free equilibrium. By applying the next generation method to find $R_{c}$, we must solve the following equation: $R_{c}=\rho\left( FV^{-1} \right)$(8). Where F is the derivatives of the new infections, V is the transition matrix (flow between compartments) and $\rho$ is the spectral radius. The entire derivation of $R_{c}$ is as follows:

$$\mathcal{F=}\left[ \begin{matrix} \left( \frac{\beta_{1}I}{N} \right)S+\left( \frac{\beta_{1}A}{N} \right)S \\ 0 \\ \begin{matrix} 0 \\ \left( \frac{\left( 1-\varepsilon_{L1} \right)\beta_{1}\left( I+I_{B} \right)}{N} \right)P+\left( \frac{\left( 1-\varepsilon_{LA1} \right)\beta_{2}\left( A+A_{B} \right)}{N} \right)P+\left( \frac{\left( 1-\varepsilon_{L2} \right)\beta_{1}\left( I+I_{B} \right)}{N} \right)F+\left( \frac{\left( 1-\varepsilon_{LA2} \right)\beta_{2}\left( A+A_{B} \right)}{N} \right)F \\ \begin{matrix} 0 \\ 0 \end{matrix} \end{matrix} \end{matrix} \right].$$

The derivative of $\mathcal{F}$ at $x_{DF}$ is

$$F=\left[ \begin{matrix} 0 & \frac{\beta_{1}S^{*}}{N^{*}} & \frac{\beta_{1}S^{*}}{N^{*}} \\ 0 & 0 & 0 \\ \begin{matrix} 0 \\ 0 \\ \begin{matrix} 0 \\ 0 \end{matrix} \end{matrix} & \begin{matrix} 0 \\ \frac{{\left( 1-\varepsilon_{L1} \right)\beta}_{1}P^{*}}{N^{*}}+\frac{{\left( 1-\varepsilon_{L2} \right)\beta}_{1}F^{*}}{N^{*}} \\ \begin{matrix} 0 \\ 0 \end{matrix} \end{matrix} & \begin{matrix} 0 \\ \frac{{\left( 1-\varepsilon_{LA1} \right)\beta}_{2}P^{*}}{N^{*}}+\frac{{\left( 1-\varepsilon_{LA2} \right)\beta}_{2}F^{*}}{N^{*}} \\ \begin{matrix} 0 \\ 0 \end{matrix} \end{matrix} \end{matrix}\begin{matrix} 0 & \frac{\beta_{1}S^{*}}{N^{*}} & \frac{\beta_{2}S^{*}}{N^{*}} \\ 0 & 0 & 0 \\ \begin{matrix} 0 \\ 0 \\ \begin{matrix} 0 \\ 0 \end{matrix} \end{matrix} & \begin{matrix} 0 \\ \frac{{\left( 1-\varepsilon_{L1} \right)\beta}_{1}P^{*}}{N^{*}}+\frac{{\left( 1-\varepsilon_{L2} \right)\beta}_{1}F^{*}}{N^{*}} \\ \begin{matrix} 0 \\ 0 \end{matrix} \end{matrix} & \begin{matrix} 0 \\ \frac{{\left( 1-\varepsilon_{LA1} \right)\beta}_{2}P^{*}}{N^{*}}+\frac{{\left( 1-\varepsilon_{LA2} \right)\beta}_{2}F^{*}}{N^{*}} \\ \begin{matrix} 0 \\ 0 \end{matrix} \end{matrix} \end{matrix} \right].$$

The transition matrix $\mathcal{V}$ is:

$$\mathcal{V=}\left[ \begin{matrix} wE \\ {-p}_{1}wE+\left( \gamma+\delta_{1} \right)I \\ \begin{matrix} -\left( 1-p_{1} \right)wE+\gamma A \\ wE_{B} \\ \begin{matrix} -p_{2}wE_{B}+\left( \gamma+\delta_{2} \right)I_{B} \\ -\left( 1-p_{2} \right)wE_{B}+\gamma A_{B} \end{matrix} \end{matrix} \end{matrix} \right].$$

The derivative of $\mathcal{V}$ at $x_{DF}$ is:

$$V=\left[ \begin{matrix} w \\ -p_{1}w \\ \begin{matrix} {-(1-p}_{1})w \\ 0 \\ \begin{matrix} 0 \\ 0 \end{matrix} \end{matrix} \end{matrix}\begin{matrix} 0 \\ \gamma+\delta_{1} \\ \begin{matrix} 0 \\ 0 \\ \begin{matrix} 0 \\ 0 \end{matrix} \end{matrix} \end{matrix}\begin{matrix} 0 \\ 0 \\ \begin{matrix} \gamma\\ 0 \\ \begin{matrix} 0 \\ 0 \end{matrix} \end{matrix} \end{matrix}\begin{matrix} 0 \\ 0 \\ \begin{matrix} 0 \\ w \\ \begin{matrix} -p_{2}w \\ -(1-p_{2})w \end{matrix} \end{matrix} \end{matrix}\begin{matrix} 0 \\ 0 \\ \begin{matrix} 0 \\ 0 \\ \begin{matrix} \gamma+\delta_{2} \\ 0 \end{matrix} \end{matrix} \end{matrix}\begin{matrix} 0 \\ 0 \\ \begin{matrix} 0 \\ 0 \\ \begin{matrix} 0 \\ \gamma\end{matrix} \end{matrix} \end{matrix} \right].$$

The inverse of V is:

$$V^{-1}=\left[ \begin{matrix} \frac{1}{w} & 0 & 0 & 0 & 0 & 0 \\ -\frac{p_{1}}{\left( \gamma+\delta_{1} \right)} & \frac{1}{\left( \gamma+\delta_{1} \right)} & 0 & 0 & 0 & 0 \\ -\frac{\left( 1-p_{1} \right)}{\gamma} & 0 & \frac{1}{\gamma} & 0 & 0 & 0 \\ 0 & 0 & 0 & \frac{1}{w} & 0 & 0 \\ 0 & 0 & 0 & -\frac{p_{2}}{\left( \gamma+\delta_{2} \right)} & \frac{1}{i} & 0 \\ 0 & 0 & 0 & -\frac{\left( 1-p_{2} \right)}{\left( \gamma+\delta_{2} \right)} & 0 & \frac{1}{\gamma} \end{matrix} \right].$$

It follows that

$$FV^{-1}=\left[ \begin{matrix} K & \frac{\beta_{1}S^{*}}{N^{*}\left( \gamma+\delta_{1} \right)} & \frac{\beta_{2}S^{*}}{N^{*}\gamma} & A & \frac{\beta_{1}S^{*}}{N^{*}\left( \gamma+\delta_{1} \right)} & \frac{\beta_{2}S^{*}}{N^{*}\gamma} \\ 0 & 0 & 0 & 0 & 0 & 0 \\ 0 & 0 & 0 & 0 & 0 & 0 \\ R & \frac{\left( 1-\varepsilon_{L1} \right)\beta_{1}P^{*}+\left( 1-\varepsilon_{L2} \right)\beta_{1}F^{*}}{N^{*}\left( \gamma+\delta_{1} \right)} & \frac{\left( 1-\varepsilon_{LA1} \right)\beta_{2}P^{*}+\left( 1-\varepsilon_{LA2} \right)\beta_{2}F^{*}}{N^{*}\gamma} & E & \frac{\left( 1-\varepsilon_{L1} \right)\beta_{1}P^{*}+\left( 1-\varepsilon_{L2} \right)\beta_{1}F^{*}}{N^{*}\left( \gamma+\delta_{2} \right)} & \frac{\left( 1-\varepsilon_{LA1} \right)\beta_{2}P^{*}+\left( 1-\varepsilon_{LA2} \right)\beta_{2}F^{*}}{N^{*}\gamma} \\ 0 & 0 & 0 & 0 & 0 & 0 \\ 0 & 0 & 0 & 0 & 0 & 0 \end{matrix} \right].$$

Where:

$$K=\frac{p_{1}\beta_{1}S^{*}}{N^{*}\left( \gamma+\delta_{1} \right)}+\frac{{(1-p}_{1})\beta_{2}S^{*}}{N^{*}\gamma}=\frac{S^{*}}{N^{*}}\left[ \frac{p_{1}\beta_{1}}{\left( \gamma+\delta_{1} \right)}+\frac{{(1-p}_{1})\beta_{2}}{\gamma} \right],$$

$$A=\frac{p_{2}\beta_{1}S^{*}}{N^{*}\left( \gamma+\delta_{2} \right)}+\frac{{(1-p}_{2})\beta_{2}S^{*}}{N^{*}\gamma}=\frac{S^{*}}{N^{*}}\left[ \frac{p_{2}\beta_{1}}{\left( \gamma+\delta_{2} \right)}+\frac{{(1-p}_{2})\beta_{2}}{\gamma} \right],$$

$$R=\frac{\left( 1-\varepsilon_{L1} \right)p_{1}\beta_{1}P^{*}+\left( 1-\varepsilon_{L1} \right)p_{1}\beta_{1}F^{*}}{N^{*}\left( \gamma+\delta_{1} \right)}+\frac{{(1-p}_{1}){\left( 1-\varepsilon_{LA1} \right)\beta}_{2}P^{*}+{(1-p}_{1}){\left( 1-\varepsilon_{LA2} \right)\beta}_{2}F^{*}}{N^{*}\gamma}=\frac{\beta_{1}p_{1}}{\left( \gamma+\delta_{1} \right)}\left[ \frac{{(1-\varepsilon}_{L1})P^{*}}{N^{*}}+\frac{{(1-\varepsilon}_{L2})F^{*}}{N^{*}} \right]+\frac{\beta_{2}(1-p_{1})}{\gamma}\left[ \frac{{(1-\varepsilon}_{LA1})P^{*}}{N^{*}}+\frac{{(1-\varepsilon}_{LA2})F^{*}}{N^{*}} \right],$$

$$E=\frac{\left( 1-\varepsilon_{L1} \right)p_{2}\beta_{1}P^{*}+\left( 1-\varepsilon_{L1} \right)p_{2}\beta_{1}F^{*}}{N^{*}\left( \gamma+\delta_{2} \right)}+\frac{{(1-p}_{2}){\left( 1-\varepsilon_{LA1} \right)\beta}_{2}P^{*}+{(1-p}_{2}){\left( 1-\varepsilon_{LA2} \right)\beta}_{2}F^{*}}{N^{*}\gamma}=\frac{\beta_{1}p_{2}}{\left( \gamma+\delta_{2} \right)}\left[ \frac{{(1-\varepsilon}_{L1})P^{*}}{N^{*}}+\frac{{(1-\varepsilon}_{L2})F^{*}}{N^{*}} \right]+\frac{\beta_{2}(1-p_{2})}{\gamma}\left[ \frac{{(1-\varepsilon}_{LA1})P^{*}}{N^{*}}+\frac{{(1-\varepsilon}_{LA2})F^{*}}{N^{*}} \right].$$

Hence the control reproduction number $R_{c}$ of model (S12) is:

$$R_{c}=\frac{S^{*}}{N^{*}}\left[ \frac{p_{1}\beta_{1}}{\left( \gamma+\delta_{1} \right)}+\frac{{(1-p}_{1})\beta_{2}}{\gamma} \right]+\frac{\beta_{1}p_{2}}{\left( \gamma+\delta_{2} \right)}\left[ \frac{{(1-\varepsilon}_{L1})P^{*}}{N^{*}}+\frac{{(1-\varepsilon}_{L2})F^{*}}{N^{*}} \right]+\frac{\beta_{2}(1-p_{2})}{\gamma}\left[ \frac{{(1-\varepsilon}_{LA1})P^{*}}{N^{*}}+\frac{{(1-\varepsilon}_{LA2})F^{*}}{N^{*}} \right] \left( S13 \right).$$

**Control Reproduction Number for the delta Variant**

Equation S12 represent the reproduction number for the delta variant, and it is in terms of the susceptible population, one dose vaccinated individuals and two dose vaccinated individuals. One possible disease-free equilibrium is where we have suspectable individuals, a proportion of individuals who have received one dose (partial immunity) and a proportion of individuals with two dose vaccination (full immunity). Consequently a scenario of the disease-free equilibrium mentioned above is $x_{DF}=\left( S^{*},0,0,0,P^{*},F^{*},0,0,0,0,0 \right)$ and the total population is $N^{*}=S^{*}+P^{*}+F^{*}. (S14)$. We can rearrange equation S13 in the following manner:

Because we want to define a proportion of susceptible, vaccinated one dose and vaccinated two dose individuals, we can divide equation20 by the total population. So,

$$\frac{S^{*}}{N^{*}}+\frac{P^{*}}{N^{*}}+\frac{F^{*}}{N^{*}}=1,$$

$$\frac{P^{*}}{N^{*}}=x and \frac{F^{*}}{N^{*}}=y.$$

It follows that,

$$\frac{S^{*}}{N^{*}}=1-x-y.$$

We rewrite equation 12 with the expression mentioned above. Hence,

$$R_{c}=\left( 1-x-y \right)\left[ \frac{p_{1}\beta_{1}}{\left( \gamma+\delta_{1} \right)}+\frac{{(1-p}_{1})\beta_{2}}{\gamma} \right]+\frac{\beta_{1}p_{2}}{\left( \gamma+\delta_{2} \right)}\left[ {(1-\varepsilon}_{L1})x+{(1-\varepsilon}_{L2})y \right]+\frac{\beta_{2}(1-p_{2})}{\gamma}\left[ {(1-\varepsilon}_{LA1})x+{(1-\varepsilon}_{LA2})y \right] \left( S15 \right).$$

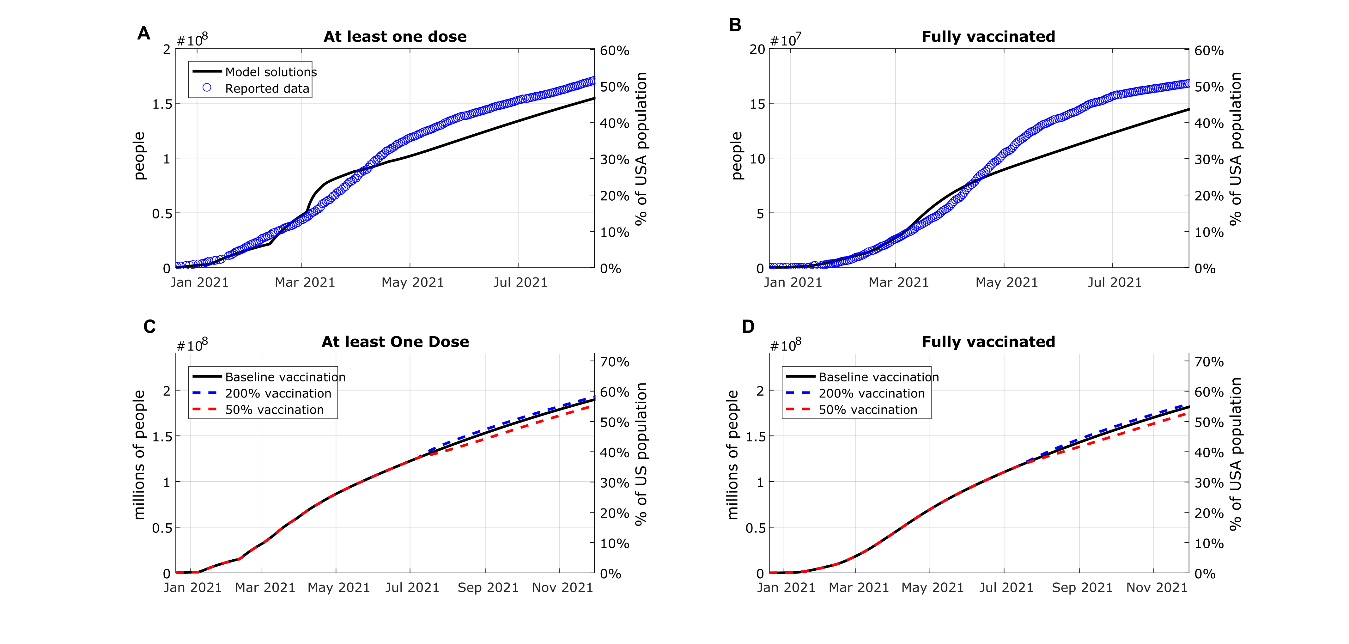
**Fig. S1.** **Fitting and projecting the vaccination rate**. (A) Deriving a function that describes the behavior of the daily doses applied in the US for one dose-vaccinated individuals. (B) Deriving a function that describes the behavior of the second dose application for individuals with one dose of vaccination. (C) Dynamics of individuals with at least one dose of vaccination considering the baseline vaccination rate since the modelled date, as well as situations where the vaccination rate is doubled or diminished 50%. (D) Dynamics of individuals with at two doses of vaccination considering the baseline vaccination rate since the modelled date, as well as situations where the vaccination rate is doubled or diminished 50%.

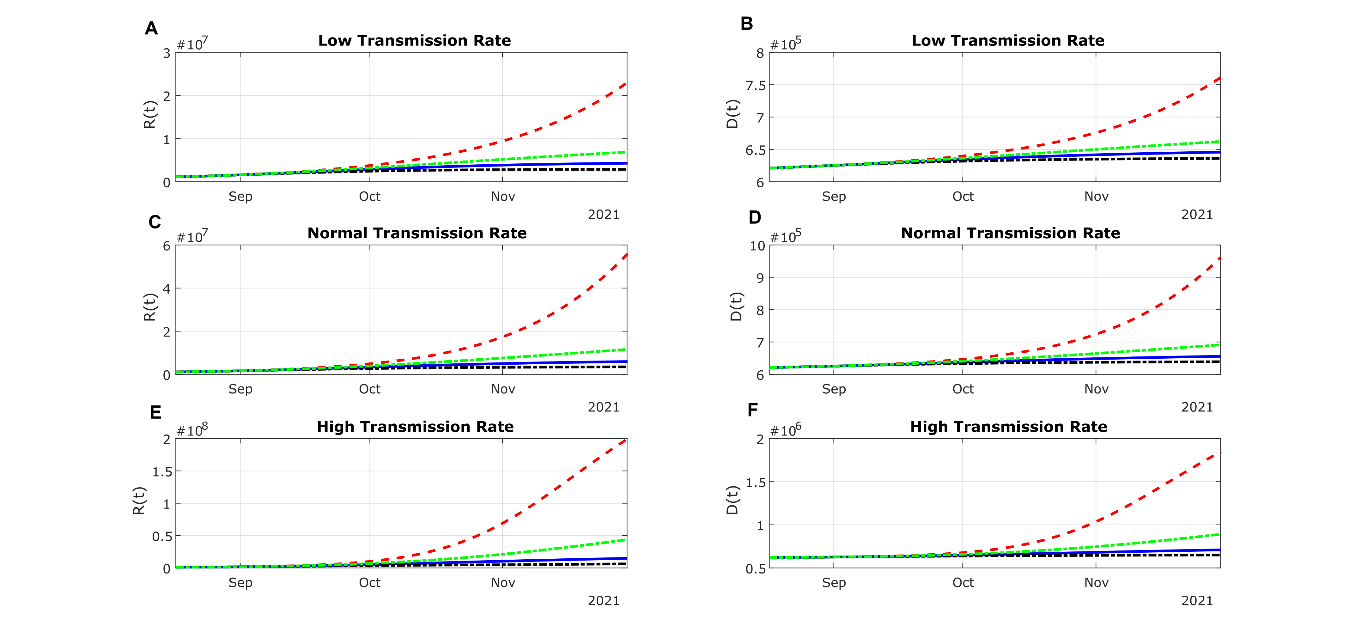

Fig. S2. Projection of the cumulative counts of recuperated and deceased individuals based on different transmission rates and vaccination rates, including (A) recuperated individuals and (B) deceased individuals under a low transmission rate; (C) recuperated individuals and (D) deceased individuals under a baseline transmission rate; (E) recuperated individuals and (F) deceased individuals under a high transmission rate. The red line presents zero vaccination; the green line represents a 50% decrease in VR; the blue dotted line means baseline VR and the black dotted line denotes 200% VR.

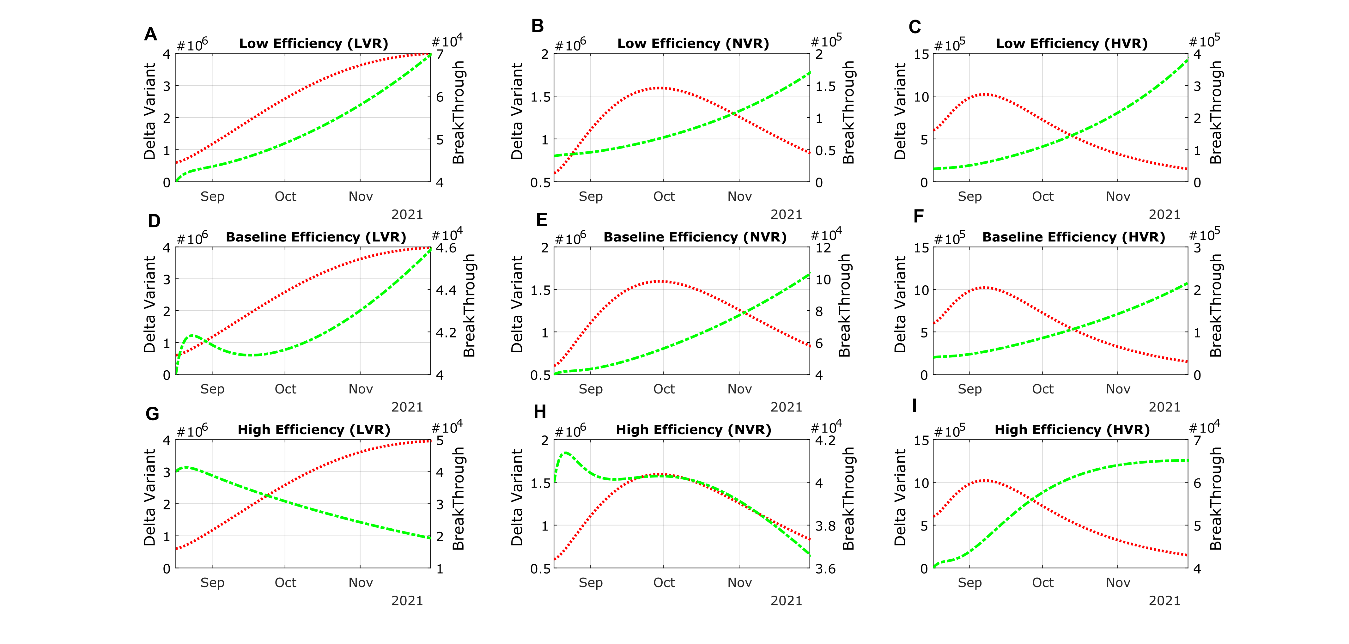

Fig. S3. Modelled projections of asymptomatic infections in unvaccinated and vaccinated subpopulations under different vaccination rates and vaccine effectiveness. The low and high vaccination effectiveness denotes the lower bound and the higher bound of the 95% confidence intervals of the real-world data, respectively. Panels (A), (D), (G) represents scenarios with a low (50%) vaccination rate, (B), (E), (H) baseline vaccination rate, and (C), (F), (I) high (200%) vaccination rate. The case counts of the unvaccinated individuals are depicted by the red line and the unit labels on the left-side y-axis, whereas the infected, vaccinated (breakthrough) cases are depicted by the green line and the unit labels on the right-side y-axis.

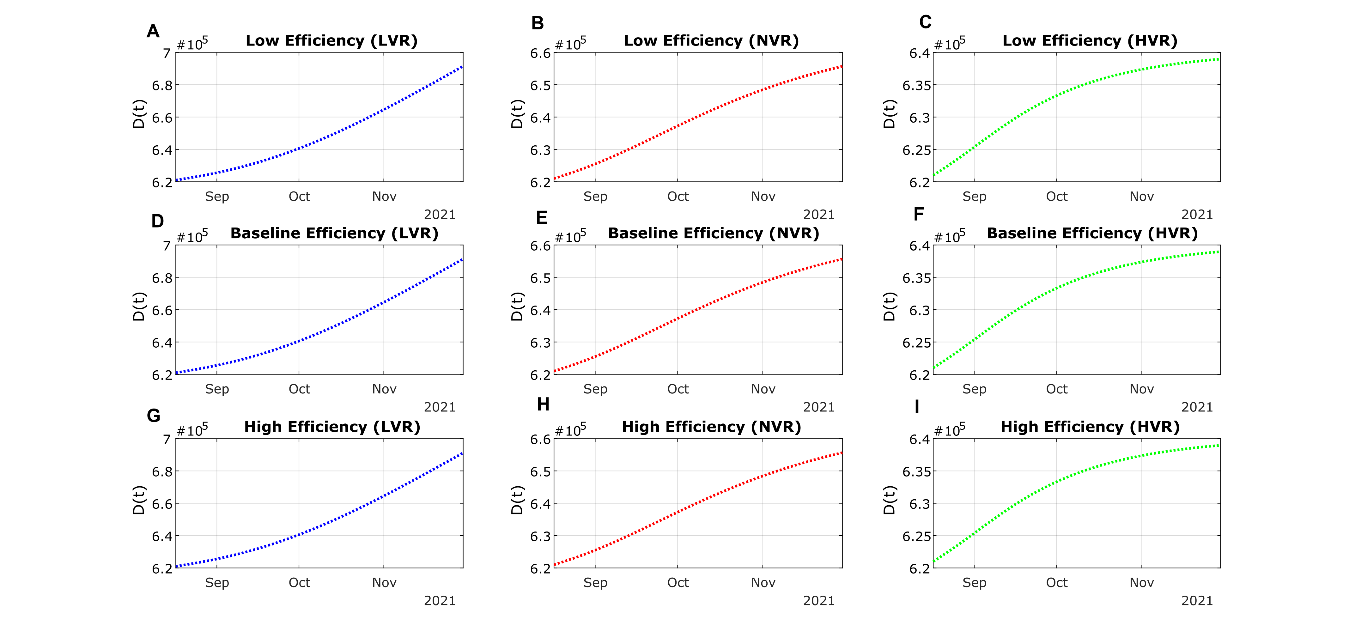

**Fig. S4.** Modelled projections of cumulative death in the US population under different vaccination rates and vaccine effectiveness. The low and high vaccination effectiveness denotes the lower bound and the higher bound of the 95% confidence intervals of the real-world data, respectively. Panels (A), (D), (G) represents scenarios with a low (50%) vaccination rate, (B), (E), (H) baseline vaccination rate, and (C), (F), (I) high (200%) vaccination rate. The case counts of the unvaccinated individuals are depicted by the red line and the unit labels on the left-side y-axis, whereas the infected, vaccinated (breakthrough) cases are depicted by the green line and the unit labels on the right-side y-axis.

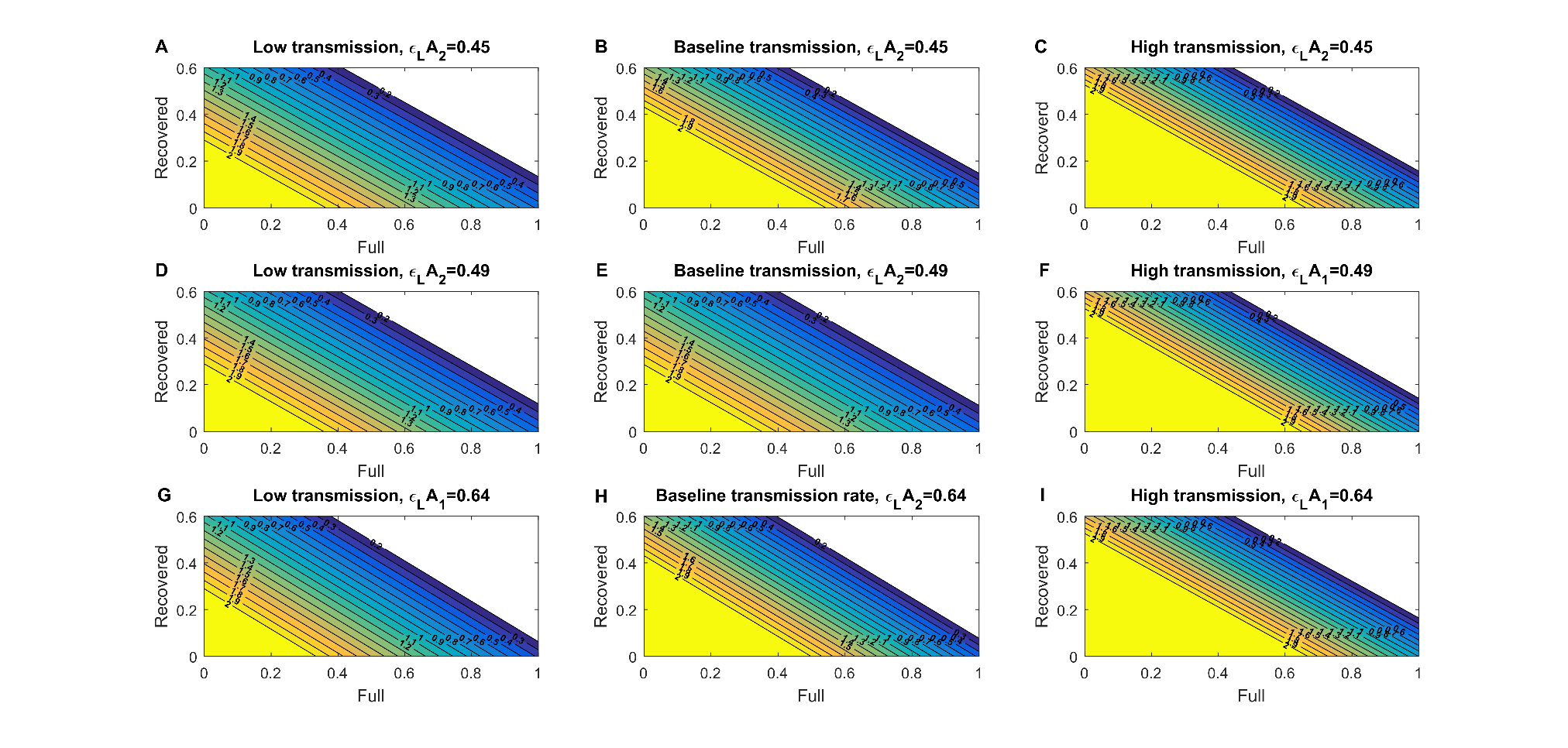

Fig. S5. Reproduction Control Number of the delta variant considering full immunity and recovered individuals for asymptomatic infections. The assumed vaccine effectiveness ($\boldsymbol{\varepsilon}_{\boldsymbol{LA}\boldsymbol{2}}$) parameter under each scenario is denoted on top of each panel and are inferred based on real-world data of individuals receiving the BNT162b2 vaccine. The heatmaps in left row consider a low transmission rate; the heatmaps in the center row panels consider a baseline transmission rate; and the heatmaps in the right row panels consider a high transmission rate.

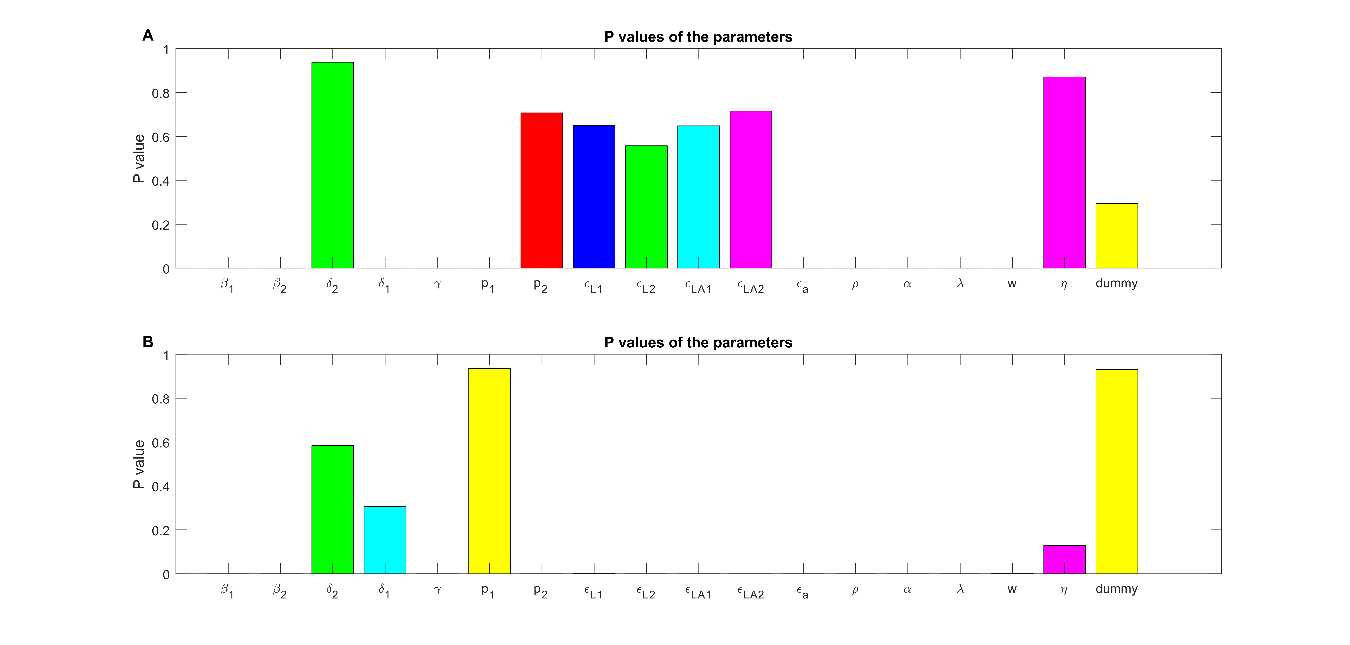

Fig. S6. Significance levels of the Partial Rank Correlation Coefficient (PRCC, depicted in Fig 6) for each parameter from the global sensitivity analyses based on our model, including the results for the (A) unvaccinated, and (B) vaccinated infected, symptomatic individuals. Visible bars where P > 0.05 denotes that the parameter was not significantly correlated with the said sub-population in the mathematical model.

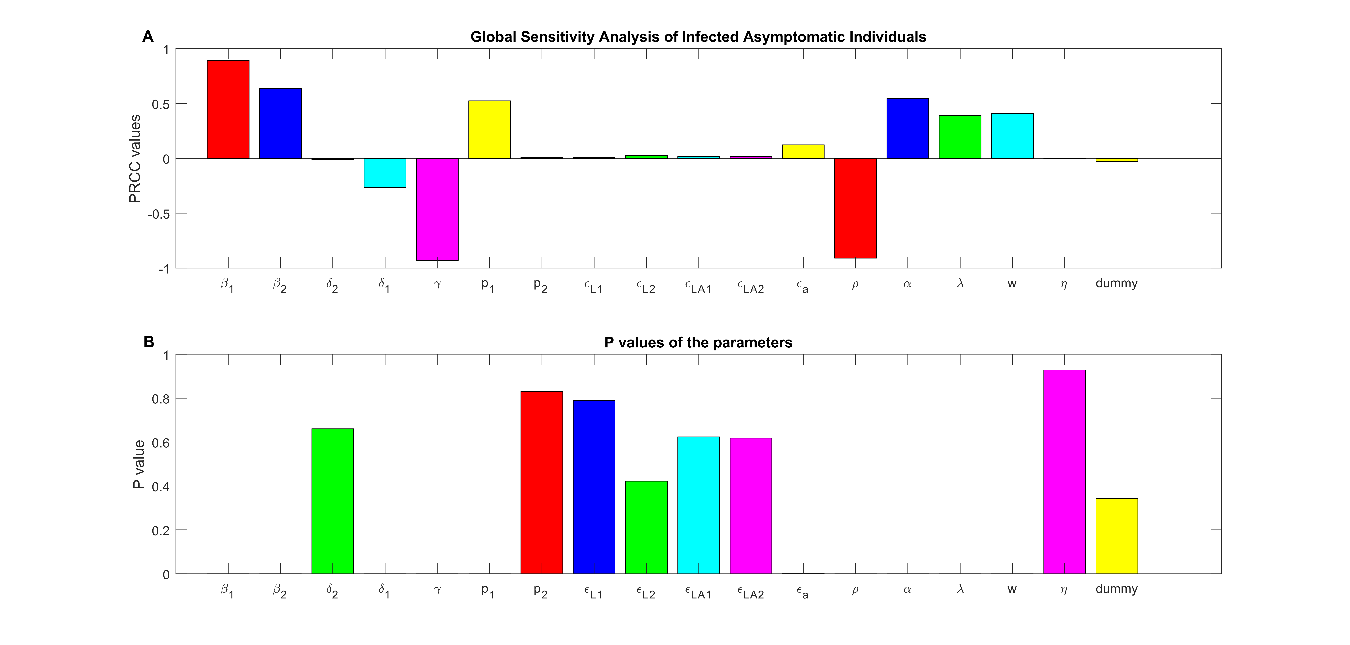

Fig. S7. Global sensitivity analyses of the parameters correlated with the unvaccinated, infected asymptomatic individuals in the mathematical model, including the (A) PRCC and (B) P value of each parameter. In the top panel, a positive PRCC indicates an increase in the parameters is correlated with an increase of the said sub-population, and a negative PRCC vice versa. In the bottom panel, parameters showing visible bars where P > 0.05 denotes that it was not significantly correlated with the said sub-population in the mathematical model.

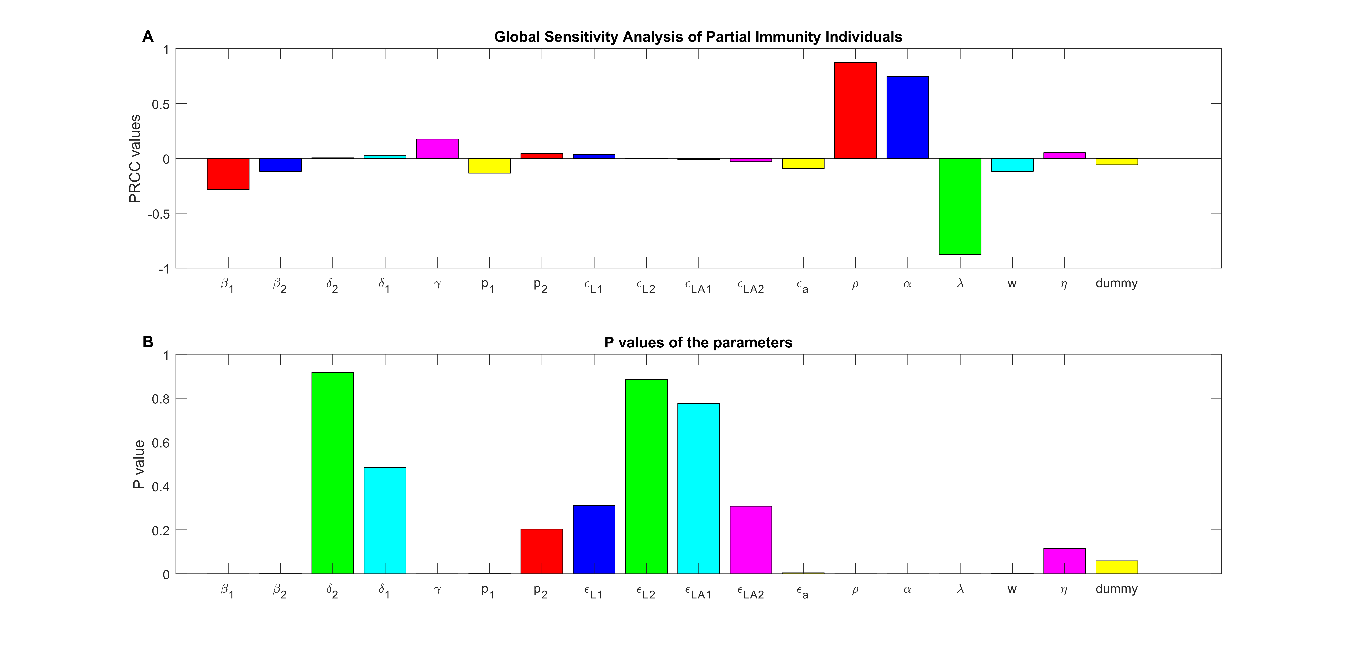

Fig. S8. Global sensitivity analyses of the parameters correlated with the partially vaccinated individuals (one dose) in the mathematical model, including the (A) PRCC and (B) P value of each parameter. In the top panel, a positive PRCC indicates an increase in the parameters is correlated with an increase of the said sub-population, and a negative PRCC vice versa. In the bottom panel, parameters showing visible bars where P > 0.05 denotes that it was not significantly correlated with the said sub-population in the mathematical model.

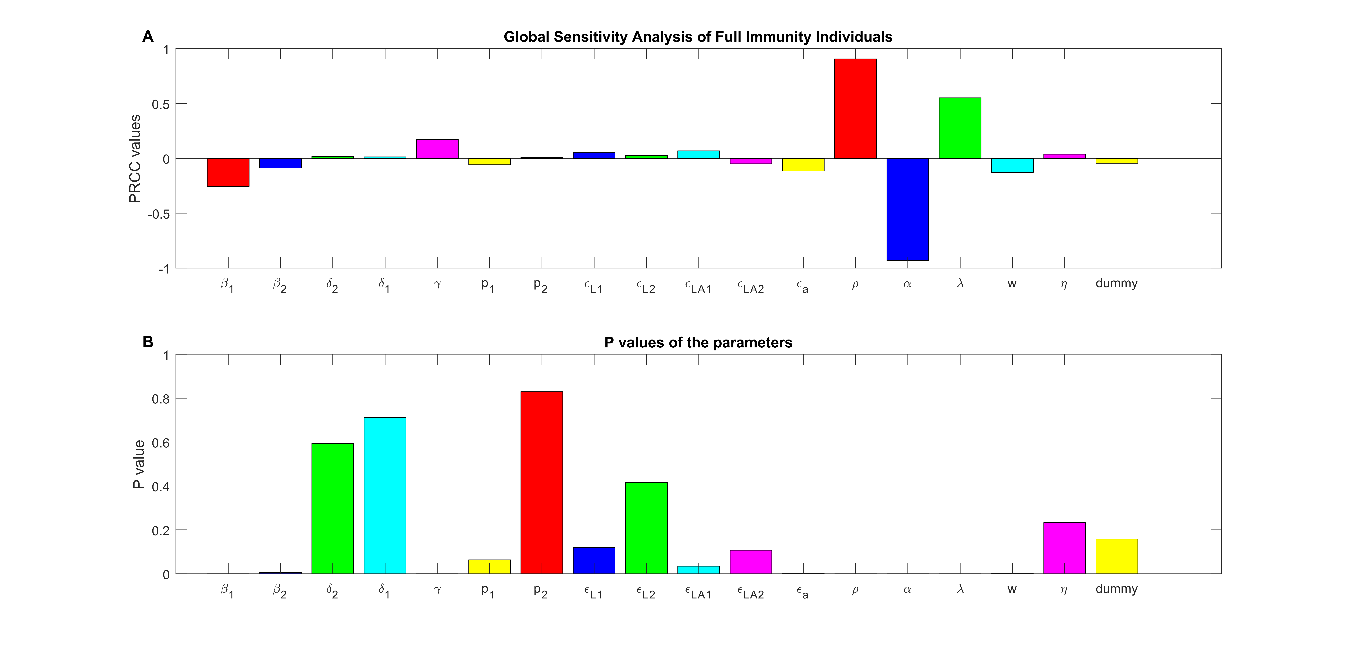

Fig. S9. Global sensitivity analyses of the parameters correlated with the fully vaccinated individuals (two doses) in the mathematical model, including the (A) PRCC and (B) P value of each parameter. In the top panel, a positive PRCC indicates an increase in the parameters is correlated with an increase of the said sub-population, and a negative PRCC vice versa. In the bottom panel, parameters showing visible bars where P > 0.05 denotes that it was not significantly correlated with the said sub-population in the mathematical model.

| Parameter | Meaning | Value | Source |
| --- | --- | --- | --- |
| $\varepsilon_{a}$ | All or nothing of the vaccine | 0.0862 | (9) |
| $\rho$ | Vaccination rate | Variable | Fitted from data |
| $\beta_{1}$ | Symptomatic infection rate | 0.5 | Fitted from data |
| $\beta_{2}$ | Asymptomatic infection rate | 0.08 | Fitted from data |
| $\alpha$ | Waning rate of the vaccine | 0.05555 | Approximated |
| $\varepsilon_{L1}$ | Vaccine effectiveness for one dose against symptomatic infection | 0.35 | (9-11) |
| $\varepsilon_{L2}$ | Vaccine effectiveness for two doses for symptomatic infection | 0.59 | (9-11) |
| $\varepsilon_{LA1}$ | Vaccine effectiveness for one dose against asymptomatic infection | 0.30 | (9-11) |
| $\varepsilon_{LA2}$ | Vaccine effectiveness for two doses against asymptomatic infection | 0.49 | (9-11) |
| $w$ | Latent period for individuals infected with delta variant | 0.25 | (*10-11*) |
| $\gamma$ | Recovery rate of individuals | 0.037 | Fitted from data |
| $\delta_{1}$ | Death rate from unvaccinated individuals | $1.8256*{10}^{-3}$ | Fitted from data |
| $\delta_{2}$ | Death rate from vaccinated individuals | $1.8256*{10}^{-5}$ | Approximated |
| $p_{1}$ | Proportion of symptomatic individuals of delta variant | 0.12 | (2) |
| $p_{2}$ | Proportion of symptomatic vaccinated individuals. | 0.09 | Approximated (11) |
| $\eta$ | Recovered individuals that get infected with the delta variant | 0.0027 | Approximated |
| $\lambda$ | Time between first and second dose | 0.0476 | (12) |

Table S1. Parameters used in the simulation of our mathematical model. Some parameters were fitted from data and the other were directly retrieved from the literature, including real-world studies and clinical trials.

**References and Notes**

1.	D. Caccavo, Chinese and Italian COVID-19 outbreaks can be correctly described by a modified SIRD model, , doi:[10.1101/2020.03.19.20039388](http://dx.doi.org/10.1101/2020.03.19.20039388).

2.	U. Avila-Ponce de León, Á. G. C. Pérez, E. Avila-Vales, An SEIARD epidemic model for COVID-19 in Mexico: Mathematical analysis and state-level forecast. *Chaos Solitons Fractals*. **140**, 110165 (2020).

3.	E. Dong, H. Du, L. Gardner, An interactive web-based dashboard to track COVID-19 in real time. *Lancet Infect. Dis.* **20**, 533–534 (2020).

4.	E. Mathieu, H. Ritchie, E. Ortiz-Ospina, M. Roser, J. Hasell, C. Appel, C. Giattino, L. Rodés-Guirao, A global database of COVID-19 vaccinations. *Nat Hum Behav*. **5**, 947–953 (2021).

5.	Á. G. C. Pérez, D. A. Oluyori, An extended SEIARD model for COVID-19 vaccination in Mexico: analysis and forecast, , doi:[10.1101/2021.04.06.21255039](http://dx.doi.org/10.1101/2021.04.06.21255039).

6.	H. S. Rodrigues, M. T. T. Monteiro, D. F. M. Torres, Sensitivity analysis in a dengue epidemiological model. *Conf. Pap. Math.* **2013**, 1–7 (2013).

7.	S. Marino, I. B. Hogue, C. J. Ray, D. E. Kirschner, A methodology for performing global uncertainty and sensitivity analysis in systems biology. *J. Theor. Biol.* **254**, 178–196 (2008).

8.	O. Diekmann, J. A. P. Heesterbeek, M. G. Roberts, The construction of next-generation matrices for compartmental epidemic models. *J. R. Soc. Interface*. **7**, 873–885 (2010).

9. J. L. Bernal, N. Andrews, C. Gower, E. Gallagher, R. Simmons, S. Thelwall, J. Stowe, E. Tessier, N. Groves, G. Dabrera, R. Myers, C. N. J. Campbell, G. Amirthalingam, M. Edmunds, M. Zambon, K. E. Brown, S. Hopkins, M. Chand, M. Ramsay, Effectiveness of Covid-19 Vaccines against the B.1.617.2 (Delta) Variant. *New England Journal of Medicine* (2021), , doi:[10.1056/nejmoa2108891](http://dx.doi.org/10.1056/nejmoa2108891).

10. Q. Li, X. Guan, P. Wu, X. Wang, L. Zhou, Y. Tong, R. Ren, K. S. M. Leung, E. H. Y. Lau, J. Y. Wong, X. Xing, N. Xiang, Y. Wu, C. Li, Q. Chen, D. Li, T. Liu, J. Zhao, M. Liu, W. Tu, C. Chen, L. Jin, R. Yang, Q. Wang, S. Zhou, R. Wang, H. Liu, Y. Luo, Y. Liu, G. Shao, H. Li, Z. Tao, Y. Yang, Z. Deng, B. Liu, Z. Ma, Y. Zhang, G. Shi, T. T. Y. Lam, J. T. Wu, G. F. Gao, B. J. Cowling, B. Yang, G. M. Leung, Z. Feng, Early Transmission Dynamics in Wuhan, China, of Novel Coronavirus–Infected Pneumonia. *N. Engl. J. Med.* **382**, 1199–1207 (2020).

11. P. Elliott, D. Haw, H. Wang, O. Eales, C. Walters, K. Ainslie, C. Atchison, C. Fronterre, P. Diggle, A. Page, Others, REACT-1 round 13 final report: exponential growth, high prevalence of SARS-CoV-2 and vaccine effectiveness associated with Delta variant in England during May to July 2021 (2021) (available at <https://spiral.imperial.ac.uk/handle/10044/1/90800>).

12. The Pfizer BioNTech (BNT162b2) COVID-19 vaccine: What you need to know, (available at <https://www.who.int/news-room/feature-stories/detail/who-can-take-the-pfizer-biontech-covid-19--vaccine>).
